# Effectiveness and safety of antiviral or antibody treatments for coronavirus

**DOI:** 10.1101/2020.03.19.20039008

**Authors:** Patricia Rios, Amruta Radhakrishnan, Jesmin Antony, Sonia Thomas, Matthew Muller, Sharon E. Straus, Andrea C. Tricco

**Affiliations:** Knowledge Translation Program, Li Ka Shing Knowledge Institute, St. Michael’s Hospital

**Keywords:** 2019-nCoV, Coronavirus, CoV – note: this one may pick up an unrelated drug name: COV155, SARS, MERS, Middle East Respiratory Syndrome, Severe Acute Respiratory Syndrome

## Abstract

**Background:** To identify safe and effective medical countermeasures (e.g., antivirals/antibodies) to address the current outbreak of a novel coronavirus (COVID-19)

**Methods:** Comprehensive literature searches were developed by an experienced librarian for MEDLINE, EMBASE, the Cochrane Library, and biorxiv.org/medrxiv.org; additional searches for ongoing trials and unpublished studies were conducted in clinicaltrials.gov and the Global Infectious Diseases and Epidemiology Network (GIDEON). Title/abstract and full-text screening, data abstraction, and risk of bias appraisal were carried out by single reviewers.

**Results:** 54 studies were included in the review: three controlled trials, 10 cohort studies, seven retrospective medical record/database studies, and 34 case reports or series. These studies included patients with severe acute respiratory syndrome (SARs, n=33), middle east respiratory syndrome (MERS, n=16), COVID-19 (n=3), and unspecified coronavirus (n=2). The most common treatment was ribavirin (n=41), followed by oseltamivir (n=10) and the combination of lopinavir/ritonavir (n=7). Additional therapies included broad spectrum antibiotics (n=30), steroids (n=39) or various interferons (n=12). No eligible studies examining monoclonal antibodies for COVID-19 were identified. One trial found that ribavirin prophylactic treatment statistically significantly reduced risk of MERS infection in people who had been exposed to the virus. Of the 21 studies reporting rates of ICU admission in hospitalized SARS or MERS patients, none reported statistically significant results in favour of or against antiviral therapies. Of the 40 studies reporting mortality rates in hospitalized SARS or MERS patients, one cohort study (MERS) and one retrospective study (SARS) found a statistically significant increase in the mortality rate for patients treated with ribavirin. Eighteen studies reported potential drug-related adverse effects including gastrointestinal symptoms, anemia, and altered liver function in patients receiving ribavirin.

**Conclusion:** The current evidence for the effectiveness and safety of antiviral therapies for coronavirus is inconclusive and suffers from a lack of well-designed prospective trials or observational studies, preventing any treatment recommendations from being made. However, it is clear that the existing body of evidence is weighted heavily towards ribavirin (41/54 studies), which has not shown conclusive evidence of effectiveness and may cause harmful adverse events so future investigations may consider focusing on other candidates for antiviral therapy.

## INTRODUCTION

### Purpose and Research Questions

The Infectious Disease Prevention and Control Branch of the Public Health Agency of Canada (PHAC) submitted a query on the effectiveness and safety of antiviral, antibody, or other medical countermeasures for the treatment of novel coronavirus (COVID-19) through the Canadian Institutes of Health Research (CIHR) Drug Safety and Effectiveness Network (DSEN). They requested the DSEN Methods and Application Group in Indirect Comparisons (MAGIC) Team conduct a rapid review on this topic with an approximate 2-week timeline.

The overall objective of this rapid review was to identify safe and effective medical countermeasures to address the current outbreak of a novel coronavirus (COVID-19). In order to focus the research question to increase feasibility, we proposed the following key research questions:

1. What is the effectiveness and safety of any antiviral and/or monoclonal antibody treatment currently available to treat (COVID-19)?
2. What is the effectiveness and safety of currently available antiviral therapies used to treat other coronavirus infections?

## METHODS

### Overall methods

The rapid review conduct was guided by the Cochrane Handbook for Systematic Reviews of Interventions^1^ along with the Rapid Review Guide for Health Policy and Systems Research^2^. The team used an integrated knowledge translation approach, with consultation from the knowledge users from the Public Health Agency of Canada at the following stages: question development, literature search, study inclusion, interpretation of results, and draft report. After the report is submitted to the Public Health Agency of Canada, a manuscript will be prepared for publication and we will invite our knowledge users to join us as coauthors. We will use the Preferred Reporting Items for Systematic Reviews and Meta-Analyses (PRISMA) Statement^3^ to guide the reporting of our rapid review results; a PRISMA extension specific to rapid reviews is currently under development.

### Literature search

Comprehensive literature searches addressing both research question 1 (RQ1) and research question 2 (RQ2) were developed by an experienced librarian for MEDLINE, EMBASE, the Cochrane Library, and biorxiv.org/medrxiv.org databases. Grey (i.e., difficult to locate or unpublished) literature was located using keyword searches of relevant terms (e.g. coronavirus, SARS, etc.) in clinicaltrials.gov and GIDEON (Global Infectious Diseases and Epidemiology Network). Additionally, the final set of included articles was cross-referenced with a list studies provided by our knowledge users from the Public Health Agency of Health as part of the scoping process for this review. The full MEDLINE search strategy and grey literature search keywords can be found in Appendix 1.

### Eligibility criteria

The Eligibility criteria followed the PICOST framework and consisted of:

#### Population (for research question 1 (RQ1) and 2 (RQ2)

Individuals of any age treated for a coronavirus infection. Subgroups of interest include older adults aged >65 years, pediatric, pregnant, or immunocompromised patients.

#### Intervention

- The following interventions were eligible for RQ1:
  - Antiviral medications approved for use in COVID-19 or currently in pre-clinical trials (animal studies, excluding *in vitro* studies) for treating COVID-19 (see Table 1).
  - Monoclonal antibodies approved for use in COVID-19 or currently in pre-clinical trials (animal studies, excluding *in vitro* studies) for treating COVID-19.
- The following interventions were of interest for RQ2:
  - Antiviral agents used alone or in combination that are approved for use in coronavirus treatment or are being examined in clinical trials for use in coronavirus treatment (Table 1).

**Table 1:**
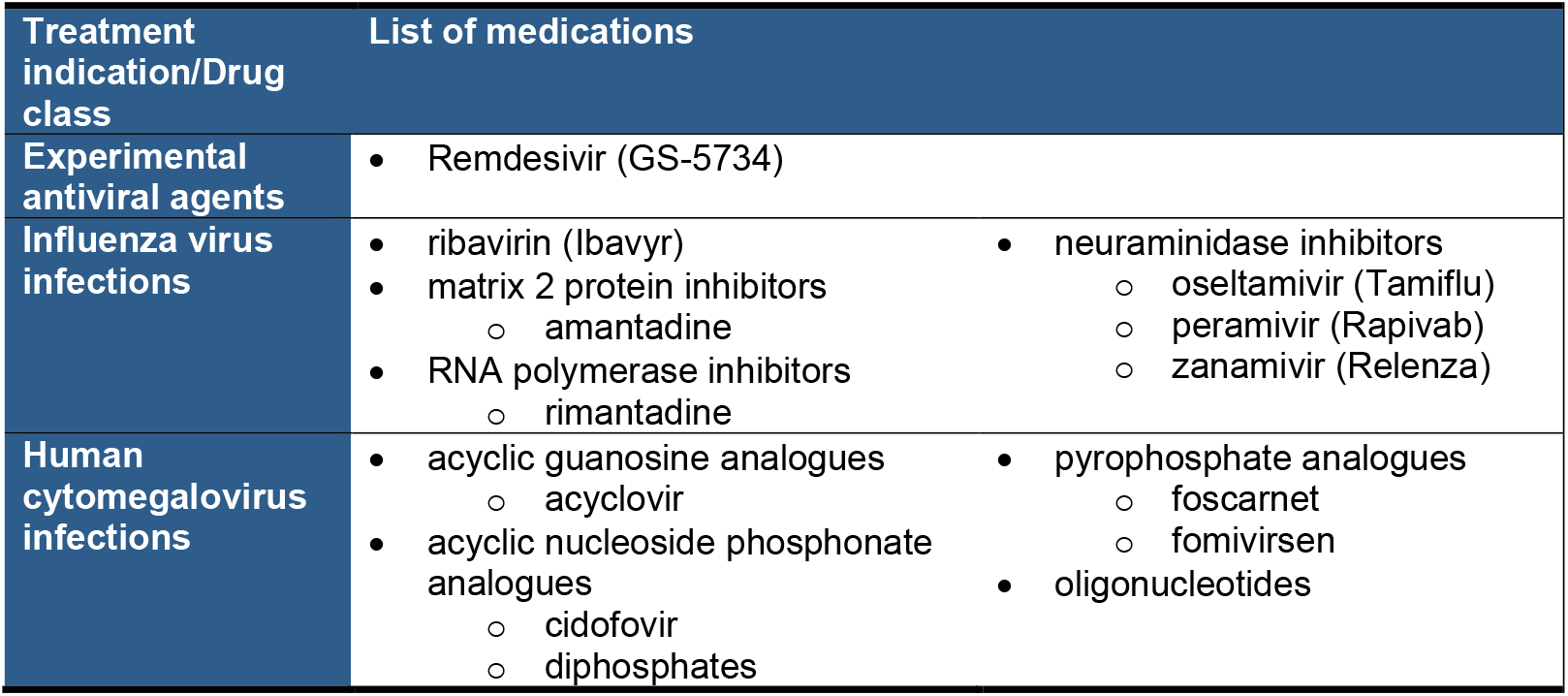
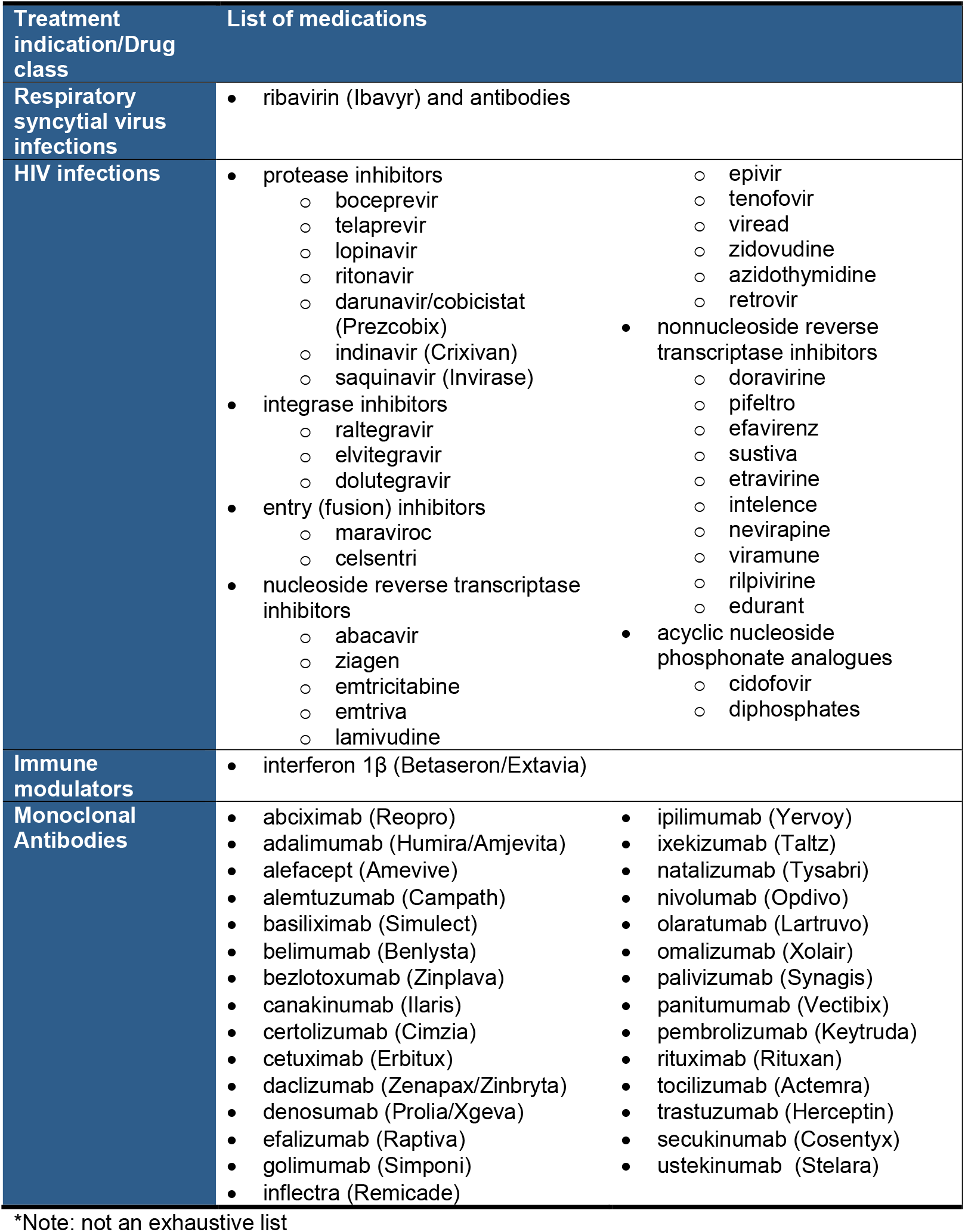
Example list of relevant interventions*.

#### Comparator (for RQ1 and RQ2)

One of the interventions listed above, no intervention, or placebo.

#### Outcomes (for RQ1 and RQ2)

lab-confirmed coronavirus infection, hospitalization, intensive care unit (ICU) admission, mortality, and adverse events (e.g., exacerbation of infection).

#### Study designs

- The following study designs were eligible for RQ1:
  - Randomized controlled trials (RCTs) and quasi-RCTs
  - Non-randomized studies (e.g., non-randomized trials, interrupted time series, controlled before after)
  - Observational studies (e.g., cohort, case control, cross-sectional)
  - Case studies, case reports, and case series
  - Pre-clinical (animal) studies
- The following study designs were eligible for RQ2:
  - Randomized controlled trials (RCTs) and quasi-RCTs
  - Non-randomized studies (e.g., quasi-RCTs, non-randomized trials, interrupted time series, controlled before after)
  - Observational studies (e.g., cohort, case control, cross-sectional)

#### Time periods (for RQ1 and RQ2)

All periods of time and duration of follow-up were eligible.

#### Other (for RQ1 and RQ2)

Only studies published in English were eligible for inclusion, due to the short timelines for this review. Relevant studies written in languages other than English and relevant studies of an ineligible design (e.g., trial protocol, literature review) will be excluded but reported in an appendix of possibly relevant articles (Appendix 2).

### Study selection

For both level 1 (title/abstract) and level 2 (full-text) screening, a screening form was prepared based on the eligibility criteria and pilot-tested by the review team using 25 citations prior to level 1 screening and 10 full text articles prior to level 2 screening. Agreement between reviewers was sufficiently high (>75%) in both cases so no further pilot-testing was required. Full screening was completed by a single reviewer for both level 1 and level 2 using Synthesi.SR, the team’s proprietary online software [https://breakthroughkt.ca/login.php].

### Data items and data abstraction

Items for data abstraction included study characteristics (e.g., study period, study design, country of conduct), patient characteristics (e.g., mean age, age range, co-morbidities), intervention details (e.g., type of intervention, dose, timing of treatment), comparator details (e.g., comparator intervention, dose), and outcome results (e.g., hospitalizations due to coronavirus, adverse events, mortality) at the longest duration of follow-up.

A standardized data abstraction form was developed to capture data on the above listed items. Prior to data abstraction, a calibration exercise was completed to test the form amongst the entire review team using two randomly selected full-text articles. Following successful completion of the calibration exercise, included studies were abstracted by single reviewers.

### Risk of bias appraisal

Risk of bias appraisal was carried out by single reviewers using Cochrane Risk of Bias (RoB) tool^4^ for controlled trials and the Newcastle Ottawa Scale^5^ (NOS) for cohort or case-control studies.

### Synthesis

Included studies were synthesized descriptively including summary statistics and detailed tables of study characteristics and results. Tables of study results are organized according to study design and where available, information on relevant subgroups were highlighted.

## RESULTS

### Literature Search

The database search returned a total of 4,491 citations, while the grey literature searches returned 305 citations, and 8 additional citations were identified from the articles provided by our knowledge users from the Public Health Agency of Canada for level 1 screening. A total of 4,567 citations were excluded after level 1 screening and a further 81 citations were identified as ineligible for the current review but potentially of interest to knowledge users, leaving 156 potentially relevant articles to be passed to level 2 screening. The full-text for 29 articles could not be obtained in time to be screened for this review and were added to the inventory of potentially relevant articles. Of the 127 articles screened at level 2, 74 were passed to data abstraction. During data abstraction a further 20 articles were excluded due to lack of relevant outcomes, leaving 54 articles included in this review (Figure 1).

**Figure 1:**
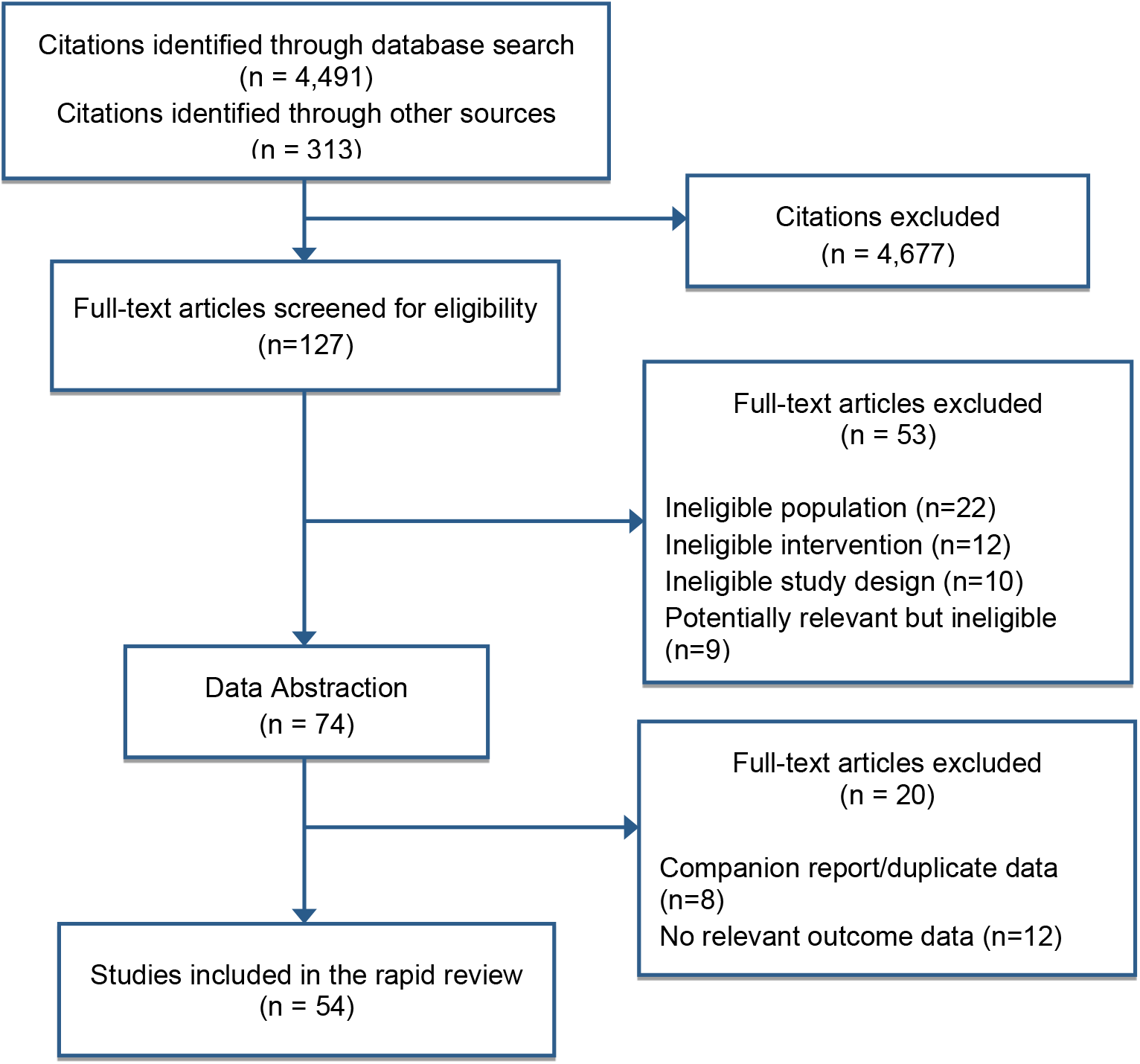
Study Flow Diagram.

### Characteristics of included studies

Of the 54 studies included in this review three were controlled trials^6-8^, 10 were cohort studies^9-18^, seven were retrospective medical record/database studies^19-25^, and 34 were case reports or case series^26-59^ (Table 2). All of the included studies were published between 2003 and 2020 with the majority conducted in Hong Kong (n=14), followed by China (n=12), Saudi Arabia (n=10), Canada (n=5), South Korea (n=4), Taiwan (n=3), and one each from France, Germany, Greece, the United Arab Emirates, and the United States. Sample sizes for the studies ranged from single patients in the case reports to groups of >1000 patients in the cohort studies. Overall, the majority of studies (n=33) dealt with treatment of Severe Acute Respiratory Syndrome (SARS), followed by Middle East Respiratory Syndrome (MERS; n=16), COVID-19 (n=3) and two studies treated unspecified coronavirus.

**Table 2:** Summary Study and Patient Characteristics.

| Characteristics (n) | Controlled Trials (n=3) | Cohort Studies (n=10) | Retrospective Studies (n=7) | Case Reports/Series (n=34) |
| --- | --- | --- | --- | --- |
| **Diagnosis** |  |  |  |  |
| COVID-19 | -- | -- | -- | 3 |
| SARS | 2 | 7 | 4 | 20 |
| MERS | 1 | 3 | 3 | 9 |
| Other coronavirus | -- | -- | -- | 2 |
| **Age of population (range)** | 22 to 57 | 15 to 70 | 22 to 79 | 4 months to 83 years |
| **Sample size [median (range)]** | 43 (16 to 190) | 169 (72 to 1934) | 63 (14 to 306) | 8 (1 to 323) |
| **Publication Year (range)** | 2004 to 2019 | 2003 to 2019 | 2003 to 2019 | 2003 to 2020 |
| **Country of conduct** | China (2), South Korea (1) | China (3), Hong Kong (3), South Korea (1), Saudi Arabia (2), Singapore (1) | Canada (2), Saudi Arabia (3), Taiwan (2) | Canada (3), China (7), France (1), Germany (1), Greece (1), Hong Kong (11), South Korea (2), Saudi Arabia (5), Taiwan (1), United Arab Emirates (1), USA (1) |
| **Comorbidities reported in study population** | No (3) | Yes (6); No (4) | Yes (5), No (2) | Yes (22), No (12) |
| **Interventions** |  | 9 |  |  |
| Ribavirin | 3 | 2 | 7 | 29 |
| Oseltamivir | -- | 2 | 1 | 7 |
| Lopinavir/ritonavir | 1 | 2 | 1 | 3 |
| *Foscarnet* | -- | -- | -- | 1 |
| *Remdesivir* | -- | -- | -- | 1 |
| *Antibiotics* | 2 | 3 | 3 | 22 |
| *Steroids* | 2 | 10 | 5 | 22 |
| *Interferons* | 1 | 3 | 2 | 6 |

The majority of studies were conducted in adult populations (n=52), one case report^31^ and one case series^36^ included infant and pediatric populations, respectively. Four case reports/series^29,45,46,48^ specifically included immunocompromised patients and one case study^34^ included a pregnant woman with MERS; however, the majority of study populations included patients with comorbid conditions (n=33). Common comorbidities included diabetes, heart disease, hypertension, and renal failure (Appendix 3). The most common antiviral studied was ribavirin (n=41), followed by oseltamivir (n=10) and the combination of lopinavir/ritonavir (n=7). Additional therapies used in the studies included a variety of broad spectrum antibiotics (n=30), steroids including hyrdcortisone, methylprednisone, or prednisolone (n=39) or various interferons (n=12; Appendix 3). No animal or human trials investigating monoclonal antibodies for the treatment of COVID-19 were found in this rapid review. All of the studies recruited from or reported on hospitalized populations and the most commonly reported outcome was mortality (n=40), followed by ICU admission (n=21) and adverse events (n=18).

### Risk of Bias Results

The 34 case reports/series and 7 retrospective studies included in this review were not assessed for risk of bias due to the inherent bias in the type of study design. The 3 trials were assessed with the Cochrane RoB tool^4^ and the 10 cohort studies were assessed using the NOS^5^. The risk of bias in the 3 included trials was overall difficult to judge due to a lack of adequate descriptions of study methods (Figure 2). All three of the trials were at high or unclear risk of bias on the following components: random sequence generation, allocation concealment, and blinding of participants/personnel (Appendix 5). The cohort studies were of fair quality overall; most of the studies suffered from a lack of representative sampling (n=8), failed to demonstrate that the outcomes of interest were not present at the start of the study (n=8), or failed to adequately ensure the comparability of cohorts (n=4; Figure 3). The complete NOS results are provided in Appendix 5.

**Figure 2.**
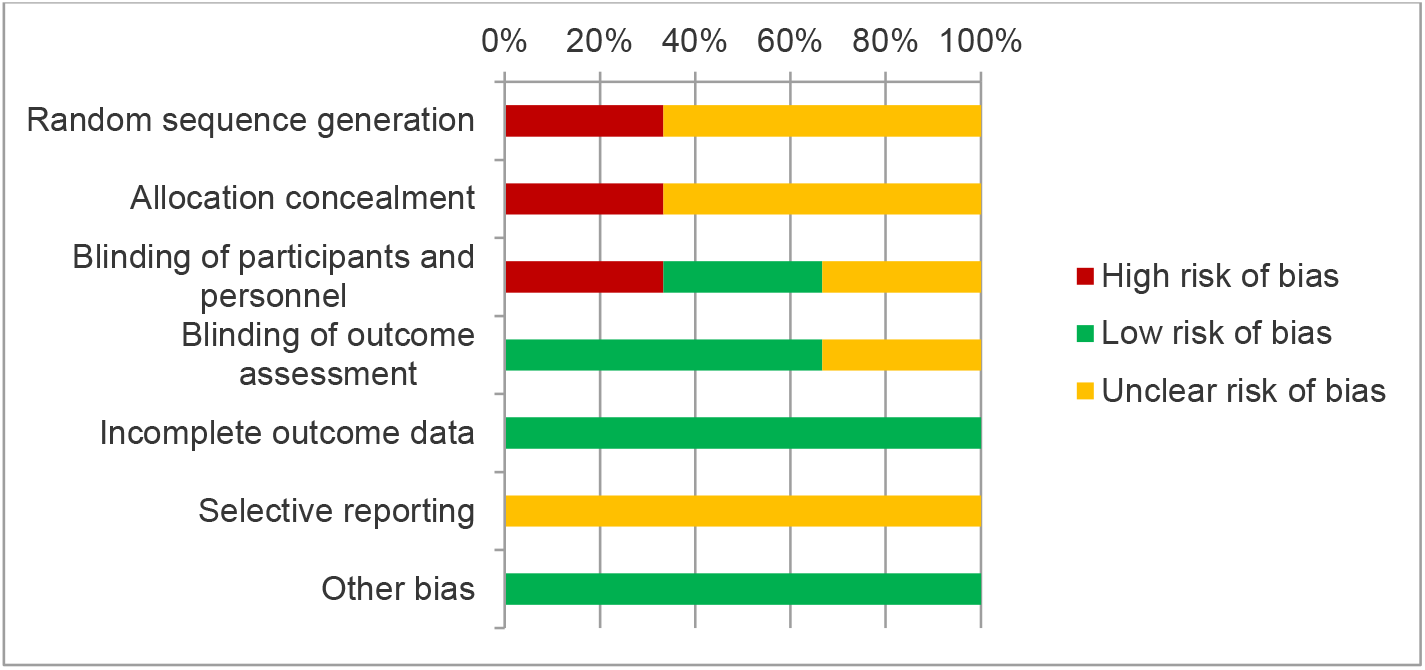
Cochrane RoB results - Controlled trials (n=3)

**Figure 3:**
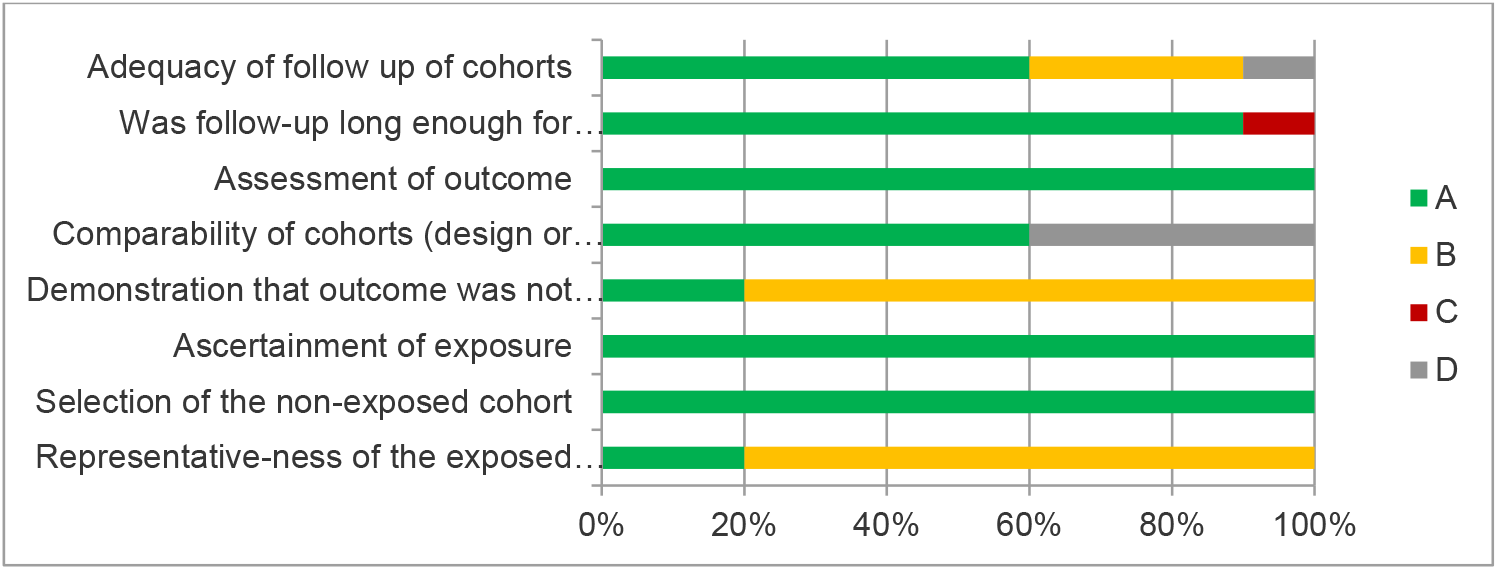
NOS Results - Cohort studies (n=10)

### Studies of COVID-19

Three studies examining patients infected with COVID-19 were included in this review: one case report^35^ and two case series^56,57^.

The case report^35^ included a 35-year-year old man, the first American diagnosed with COVID-19. He, he was initially treated with vancomycin and cefepime which are standard treatments for suspected community-acquired pneumonia. Upon lab-confirmation of COVID-19 infection, the antibiotics were stopped and the patient was started on Remdesivir 7 days after initial admission to hospital. At study end, the patient remained hospitalized with the majority of symptoms resolved (see appendices 3 and 4 for complete details).

The two case series^56,57^ were conducted in China and included 4 and 138 patients, respectively. All patients were hospitalized and initial diagnosis was made based on WHO Criteria later confirmed by lab-testing of the patient specimens. The case series included an approximately even number of male and female (55% v 45%) patients ranging in age from 19 to 68 years old, with a variety of co-morbidities including cardiovascular disease, chronic kidney or liver disease, COPD, and diabetes (Appendix 3). In one case series^57^, patients (n=4) were treated with a combination of lopinavir/ritonavir, Arbidol (umifenovir), antibiotics, Shufeng Jiedu Capsule (Traditional Chinese Medicine), and intravenous immunoglobulins (Appendix 4). At study end (15 days),) two patients tested negative for COVID-19 and were subsequently discharged from the hospital and two patients remained hospitalized, one of whom still required mechanical ventilation (Appendix 4). In the larger case series^56^, 124 patients were treated with oseltamivir combined with antibiotic therapy in 89 patients and combined with glucocorticoids in 62 patients (Appendix 4). Over the course of the study, 34 patients treated with oseltamivir were admitted to the ICU, 17 of which required invasive mechanical ventilation. At study end (19 days),) 47 patients had been discharged and 6 patients died, all of whom had been admitted to ICU (Appendix 4).

### Ongoing human trials for COVID-19

Four currently ongoing randomized controlled trials proposing to test treatments for COVID-19 were identified through keyword searches of clinicaltrials.gov. All four trials are being carried out in China, three are investigating antiviral medications (lopinavir/ritonavir, arbidol (umifenovir), darunavir, cobicstat, and, ASC09/ritonavir) and one trial is investigating a combination of lopinavir/ritonavir with Traditional Chinese Medicines (TCM). At the time of this writing two of the trials have started recruiting patients (further details in Table 3).

**Table 3:** Details of ongoing COVID-19 trials.

| Author, Year<br>Country<br>NCT ID | Status<br>Estimated Enrollment<br>Estimated completion | Eligibility Criteria (age; diagnosis)<br>Interventions |
| --- | --- | --- |
| Li, 2020<br>China<br>NCT04252885 | Recruiting<br>125 participants<br>July 31, 2020 | Adult (18-80 yrs); lab-confirmed infection<br>Group A: standard treatment + lopinavir/ritonavir<br>Group B; standard treatment + arbidol (umifenovir)<br>Group C: standard treatment |
| Lu, 2020<br>China<br>NCT04252274 | Not yet recruiting<br>30 participants<br>December 31, 2020 | All ages; National Health Commission diagnostic criteria<br>Intervention: Darunavir, Cobicistat + conventional treatments<br>Comparator: Conventional treatments |
| Qiu, 2020<br>China<br>NCT04261907 | Not yet recruiting<br>160 participants<br>June 30, 2020 | Adult (18-75 yrs); lab-confirmed infection<br>Intervention: ASC09/ritonavir + conventional treatment<br>Comparator: lopinavir/ritonavir + conventional treatment |
| Xiao, 2020<br>China<br>NCT04251871 | Recruiting<br>150 participants<br>January 22, 2021 | Youth/Adult (14-80 yrs); lab-confirmed infection<br>Intervention: TCM + conventional medicines**<br>Comparator: Conventional medicines** |

\*\*Conventional medicines includes: oxygen therapy, antiviral therapy (alfa interferon via aerosol inhalation, and lopinavir/ritonavir, 400mg/100mg, p.o, bid)

### Effectiveness Outcomes

#### Infection Prevention

One of the included trials^7^ examined the effectiveness of ribavirin combined with lopinavir/ritonavir compared to no treatment as a prophylactic measure for healthcare workers highly exposed to MERS through unprotected exposure to a patient with pneumonia later confirmed to be caused by MERS-CoV. None of the subjects in the prophylaxis arm (ribavirin/lopinavir/ritonavir) developed MERS while 6 subjects in the control arm were infected with MERS as confirmed by rPT-PCR testing. The risk of infection was statistically significantly lower in the prophylaxis arm (adjusted odds ratio: 0.405, 95% CI 0.274 to 0.599, p=0.009; Appendix 4).

#### ICU Admission

Of the 21 studies reporting this outcome, one was a randomized trial^6^ comparing ribavirin supplemented with hydrocortisone to ribavirin alone; three were cohort studies^14,15,17^ comparing oseltamivir to steroid treatment alone, ribavirin with continuous steroid treatment to ribavirin with high-dose ‘pulse’ steroids, and ribavirin to steroid and/or antibiotic treatment; three were retrospective studies^19,20,25^ examining the effectiveness of ribavirin and oseltamivir alone or in combination with other drugs; and 14 were case reports/series^33,34,37,40,42,44,45,47,50,51,53,55-57^ examining ribavirin, oseltamivir, lopinavir/ritonavir alone or in combination with steroids or antibiotics. None of the trials, cohorts, or retrospective studies demonstrated statistically significant results between any of the comparisons (i.e., in favour of or against the effectiveness of ribavirin, oseltamivir or lopinavir/ritonavir) in reducing the risk of ICU admission for patients with SARS or MERS. The case reports and series were similarly inconclusive, none of the study authors reported a particular advantage for patients with COVID-19, SARS, or MERS treated with ribavirin, oseltamivir, or lopinavir/ritonavir.

#### Special populations

One case series^45^ included 4 patients with hematological malignancies that acquired MERS infections. The patients were all treated with oseltamivir and one patient required admission to the ICU due to worsening symptoms. One case report^34^ of a pregnant woman with MERS described initially attempting treatment with antibiotics but the patient did not respond and was transferred to ICU where antiviral treatments were initiated but the patient continued to deteriorate and died. In a case series^36^ of 4 pediatric patients with SARS, all 4 were treated with ribavirin and 2 patients required mechanical ventilation during the course of their illness.

#### Mortality

Mortality was reported in two of the included trials (ribavirin), all 10 cohort studies (ribavirin, lopinavir/ritonavir, oseltamivir), all seven retrospective studies (ribavirin, lopinavir/ritonavir, oseltamivir), and 21 case reports or case series (ribavirin, oseltamivir, lopinavir/ritonavir). The comparative studies (trials and cohorts) failed to find statistically significant results indicating that none of the antivirals they examined were effective in reducing mortality for SARS or MERS. One cohort study^10^ of MERS patients found that treatment with ribavirin and interferons significantly increased 90-day mortality risk (adjusted odds ratio: 2.27, 95% CI 1.20-4.32). The patients in this cohort were generally older (median age 57 (IQR 47-70)) and had a number of underlying chronic conditions including diabetes, cardiovascular disease, chronic lung, renal, or liver disease and malignancy including leukemia or lymphoma which may in part explain the increased risk. One retrospective study^20^ of SARS patients found the 21-day mortality rate was significantly higher in a cohort of patients treated with ribavirin compared to matched historical controls (6.5%, 95% CI 1.9% to 11.8%). Patients in this study were largely middle aged (34 to 57 years of age); however a large proportion of patients that died (approximately 80%) had underlying conditions such as diabetes or cancer.

The two case series^56,57^ and one case report^35^ that included patients with COVID-19 that used Remdesivir (1 patient), lopinavir/ritonavir (4 cases) and oseltamivir (124 cases) reported 6 deaths in the cohort treated with oseltamivir.

#### Special Populations

The four cases reports/series^29,45,46,48^ that included immunosuppressed patients with MERS (5 patients) and unspecified coronavirus (2 patients) reported 3 deaths all in patients with hematological malignancies treated with foscarnet (n=1) and oseltamivir (n=2). One patient with HIV and 2 patients with hematological malignancies that acquired MERS were treated with ribavirin and oseltamivir respectively and survived after being hospitalized for their illness (38 and 28 days respectively). The case report^34^ of a pregnant woman with MERS treated with oseltamivir and later ribavirin succumbed to septic shock 8 days after admission to hospital. The two case series^31,36^ that included pediatric patients treated with ribavirin reported no mortality at study end.

### Safety Outcomes

#### Adverse Events

One of the included trials^7^, seven of the cohort studies^9-11,13-15,17^, three of the retrospective studies^20,21,25^, and seven case reports/series^28,38,39,42,43,50,51^ reported treatment related adverse events while two retrospective studies and three case reports/series reported that no treatment related adverse events occurred. In the trial^7^ examining the effectiveness of ribavirin/lopinavir/ritonavir compared to no treatment as a prophylactic measure for healthcare workers, treatment-related adverse events were widely reported in the prophylaxis arm, including: GI symptoms (diarrhea n=9, nausea n=9, stomatitis n=4), anemia (n=9), leucopenia (n=8) and hyperbilirubinemia (n=20). All adverse effects occurred during prophylactic therapy and resolved shortly after conclusion of treatment with no further intervention. Overall, the most commonly reported adverse events were anemia (n=12 studies) and altered liver function (n=5 studies) in patients treated with ribavirin. Other treatment related adverse events included gastrointestinal symptoms (e.g., nausea, vomiting), changes in kidney function, cardiac events (e.g., bradycardia, atrial fibrillation), hyperglycemia, and changes in mental status (e.g., confusion, anxiety). It should be noted however, that in the studies reporting cardiac adverse events, hyperglycemia, and mental status changes patients were receiving steroids as well as ribavirin.

#### Special Populations

None of the studies that included special populations reported treatment-related adverse events.

## DISCUSSION

The Public Health Agency of Canada commissioned a rapid review to address the urgent question of the effectiveness and safety of antiviral or antibody therapies in the treatment of coronavirus. A comprehensive literature search of both electronic databases and grey literature sources resulted in 54 studies of various antiviral treatments in patients diagnosed with COVID-19, SARS, or MERS; however, no animal or human studies of monoclonal antibodies could be found.

Overall the results of the included studies proved inconclusive on the effectiveness of antiviral drugs in treating coronavirus infections and prevent any particular treatments from being recommended for use. There is a low quality of available evidence that largely consists of case reports and case series, with few observational studies, and even fewer trials. There were however important safety signals identified in the included studies, particularly the possible development of anemia and altered liver function in patients receiving ribavirin treatment. It is similarly difficult to recommend a particular antiviral drug as a promising candidate for further investigation due to the variable quality and inconclusive results of the current evidence. This review does show however that the existing body of evidence is weighted heavily towards studies of ribavirin which has shown no particular efficacy in treating coronavirus and may in fact cause harmful adverse effects. Future investigations into potential antiviral therapies for coronavirus may be best served by pointing their attention to other drug candidates.

There are several limitations to the review methods employed here, single screening and abstraction for example, however they were selected to thoughtfully tailor our methods according to our knowledge user needs and the urgent nature of the request to provide timely results.

## CONCLUSIONS

The current evidence for the effectiveness of antiviral therapies for coronavirus is not conclusive and suffers from a lack of well-designed prospective trials or observational studies. None of the interventions examined in this review can be recommended for use in patients with coronavirus. Similarly, no firm recommendations can be made for or against these interventions from a safety perspective due to a lack of conclusive evidence. Some important safety signals potentially related to ribavirin use were identified (anemia, altered liver function) but also require further investigation to clarify their relation to the drug.

## Data Availability

All datasets supporting the conclusions of this article are included within the article.

## Acknowledgements

Jessie McGowan (literature search), Tamara Rader (literature search), Krystle Amog (report preparation), Chantal Williams (report preparation), Naveeta Ramkissoon (report preparation)

## APPENDIX 1 Search Strategies

### MEDLINE Search Strategy

1. coronaviridae infections/ or coronavirus infections/ or severe acute respiratory syndrome/ or SARS Virus/
2. (coronavirus* or corona virus* or mers or middle east respiratory syndrome* or Severe Acute Respiratory Syndrome* or SARS or CoV or SARS-CoV or MERS-CoV or 2019-nCoV).tw,kf.
3. 3 or/1-2
4. dt.fs.
5. exp Antiviral Agents/
6. (antiviral or anti-viral or anti viral).tw,kf.
7. (neuraminidase adj2 inhibitor).tw,kf.
8. Remdesivir.tw,kf.
9. (oseltamivir or Tamiflu or peramivir or Rapivab or zanamivir or Relenza or ribavirin or Ibavyr).tw,kf.
10. (matrix adj3 inhibitors).tw,kf.
11. exp DNA-Directed RNA Polymerases/
12. RNA polymerase inhibitors.tw,kf.
13. Rimantadine/
14. Rimantadine.tw,kf.
15. acyclic guanosine analogues.tw,kf.
16. Acyclovir/
17. Acyclovir.tw,kf.
18. acyclic nucleoside phosphonate analogues.tw,kf.
19. Cidofovir/
20. (diphosphate or Cidofovir).tw,kf.
21. Diphosphonates/
22. pyrophosphate analogues.tw,kf.
23. Foscarnet/
24. Foscarnet.tw,kf.
25. Oligonucleotides/
26. Fomivirsen.tw,kf.
27. Protease Inhibitors/
28. (boceprevir or telaprevir or lopinavir or ritonavir or darunavir or cobicistat or Prezcobix or indinavir or Crixivan or saquinavir or Invirase).tw,kf.
29. Integrase Inhibitors/
30. (raltegravir or elvitegravir or dolutegravir).tw,kf.
31. HIV Fusion Inhibitors/
32. (maraviroc or Celsentri).tw,kf.
33. Reverse Transcriptase Inhibitors/
34. nucleoside reverse transcriptase inhibitors.tw,kf.
35. (abacavir or Ziagen or emtricitabine or Emtriva or lamivudine or Epivir or tenofovir or Viread or zidovudine or azidothymidine or Retrovir).tw,kf.
36. nonnucleoside reverse transcriptase inhibitors.tw,kf.
37. (doravirine or Pifeltro or efavirenz or Sustiva or etravirine or Intelence or nevirapine or Viramune or rilpivirine or Edurant).tw,kf.
38. exp Interferon beta-1b/
39. (Betaseron or Extavia).tw,kf.
40. or/5-39
41. Antineoplastic Agents, Immunological/
42. (abciximab or Reopro or adalimumab or Humira or Amjevita or alefacept or Amevive or alemtuzumab or Campath or basiliximab or Simulect or belimumab or Benlysta or bezlotoxumab or Zinplava or canakinumab or Ilaris or certolizumab or Cimzia or cetuximab or Erbitux or daclizumab or Zenapax or Zinbryta or denosumab or Prolia OR, Xgeva or efalizumab or Raptiva or golimumab or Simponi or inflectra or Remicade or ipilimumab or Yervoy or ixekizumab or Taltz or natalizumab or Tysabri or nivolumab or Opdivo or olaratumab or Lartruvo or omalizumab or Xolair or palivizumab or Synagis or panitumumab or Vectibix or pembrolizumab or Keytruda or rituximab or Rituxan or tocilizumab or Actemra or trastuzumab or Herceptin or secukinumab or Cosentyx or ustekinumab or Stelara).tw,kf.
43. exp Antibodies, Monoclonal/
44. or/41-43
45. medical countermeasures/
46. (countermeasure* or counter measure*).tw,kf.
47. 45 or 46
48. 4 or 40 or 44 or 47
49. 3 and 48
50. animals/ not humans/
51. 49 not 50

**Grey Literature: ClinicalTrials.gov and GIDEON (Global Infectious Diseases and Epidemiology Network).**

## APPENDIX 2 Potentially relevant articles not included in this review

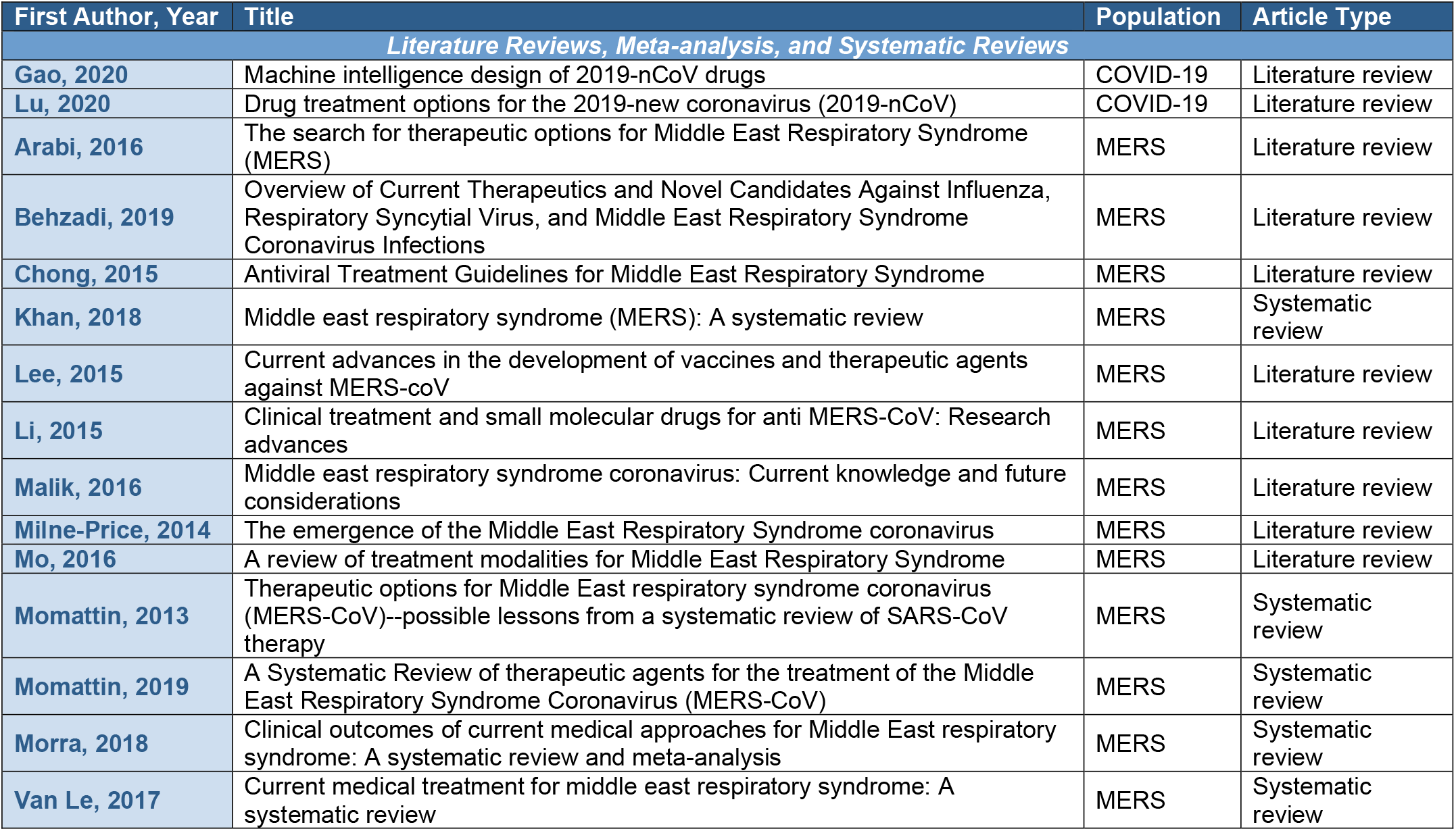
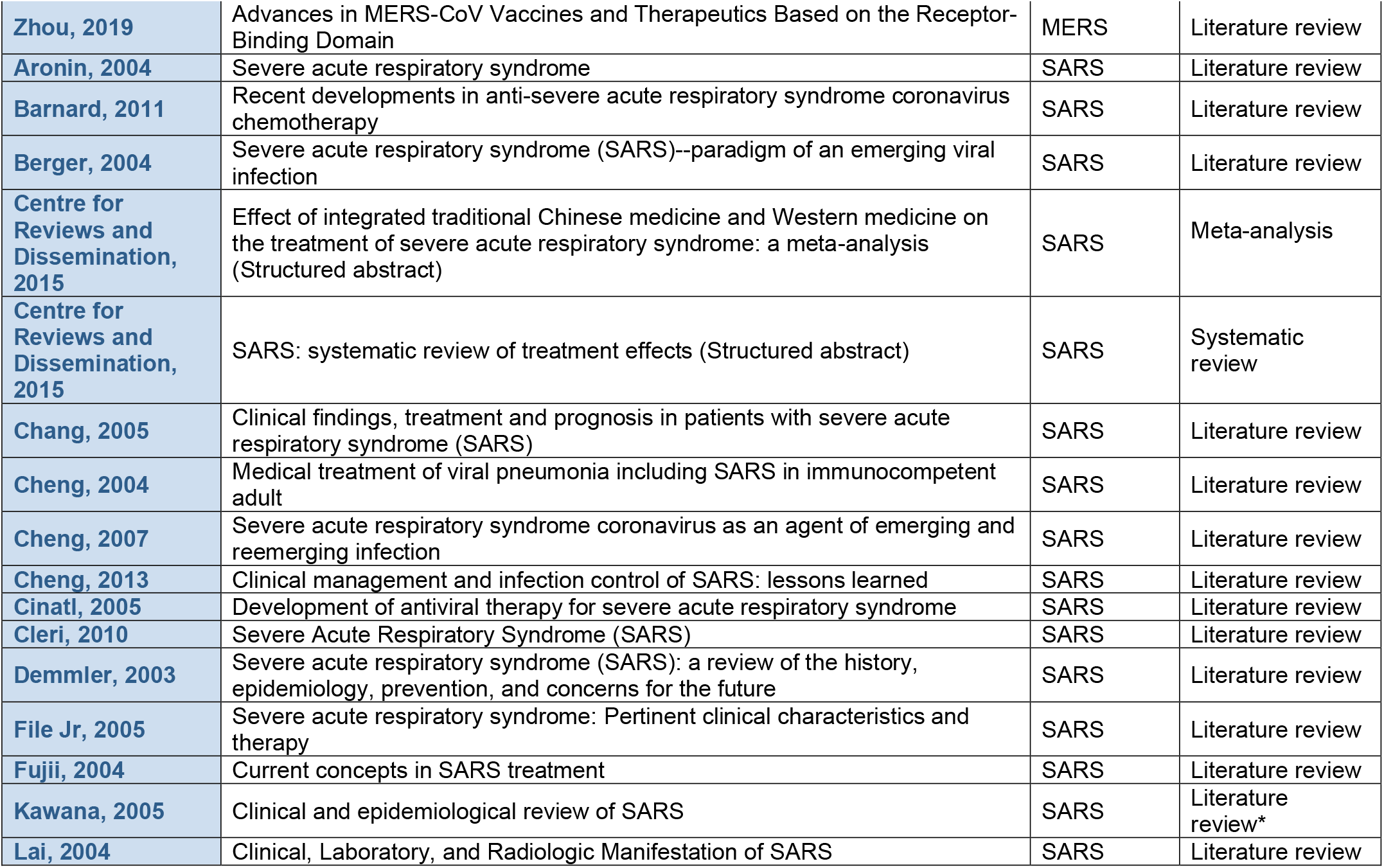
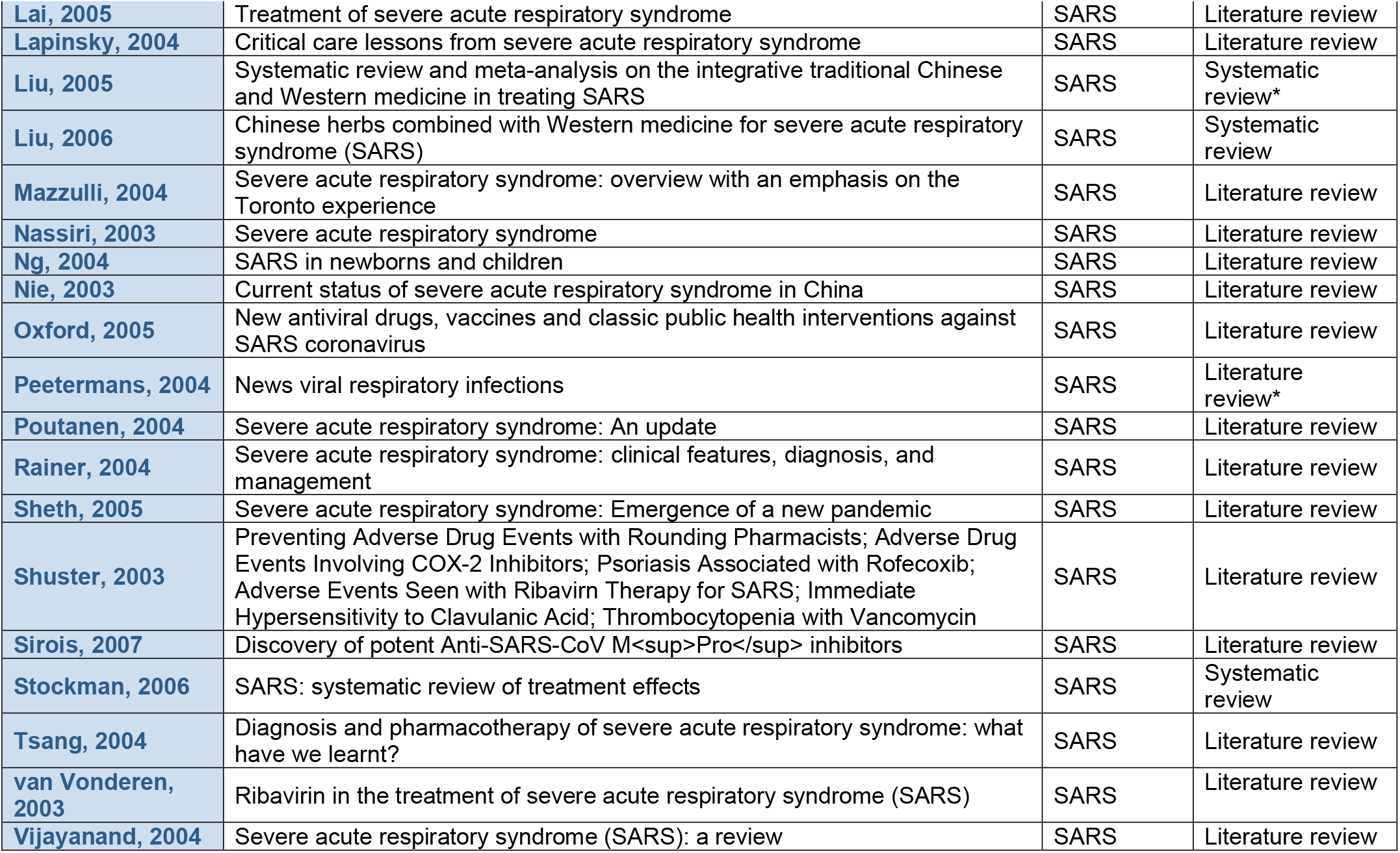
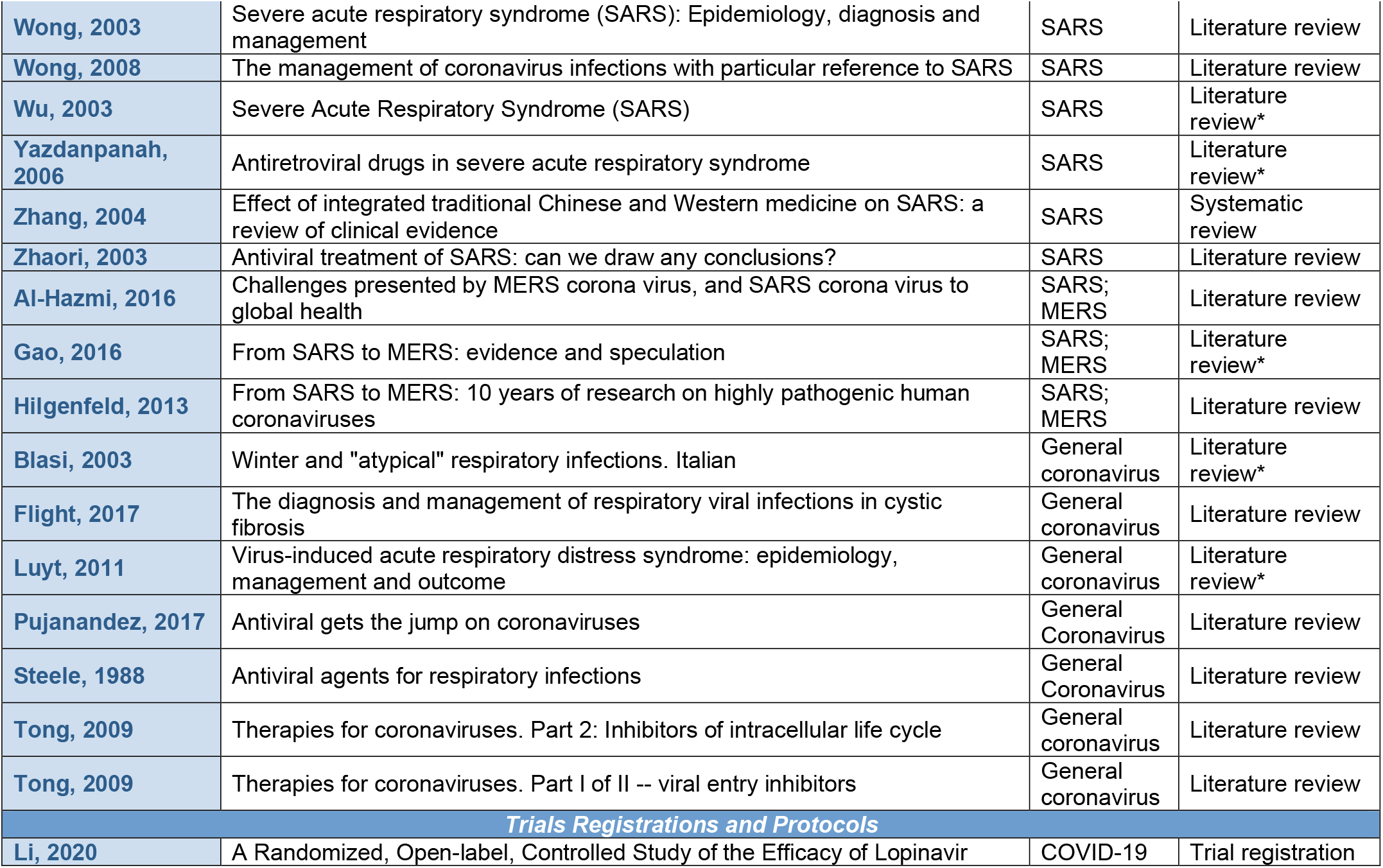
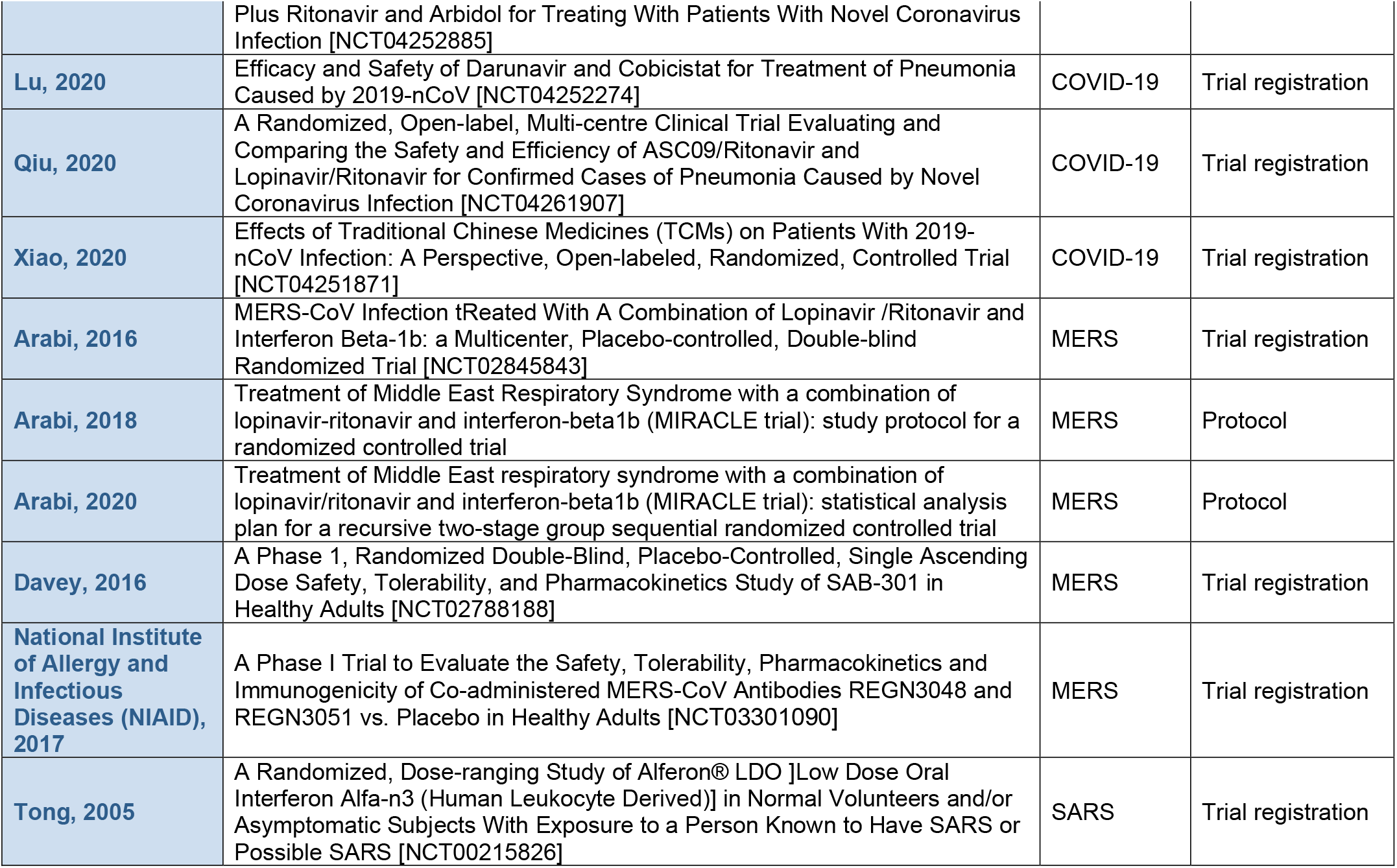
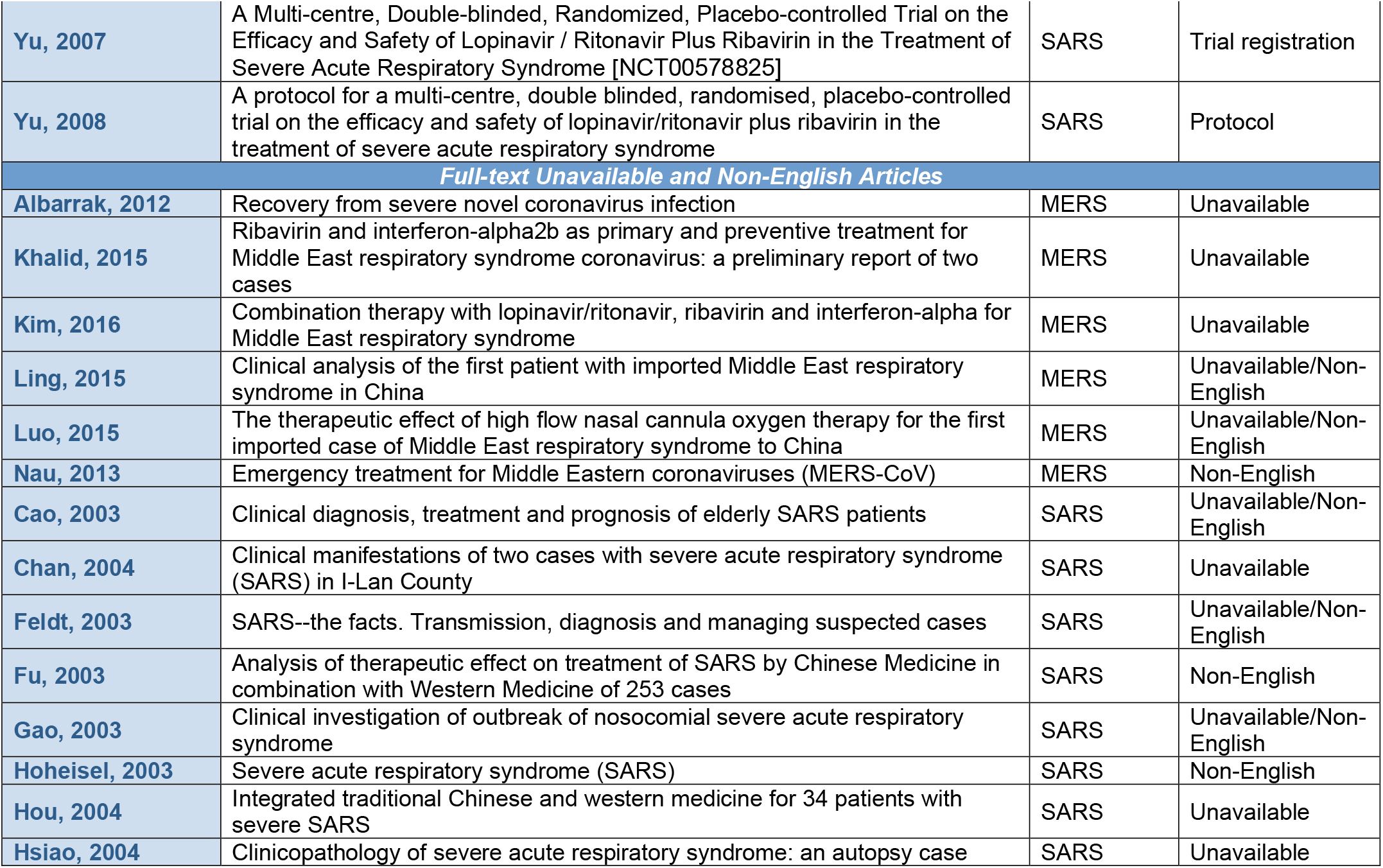
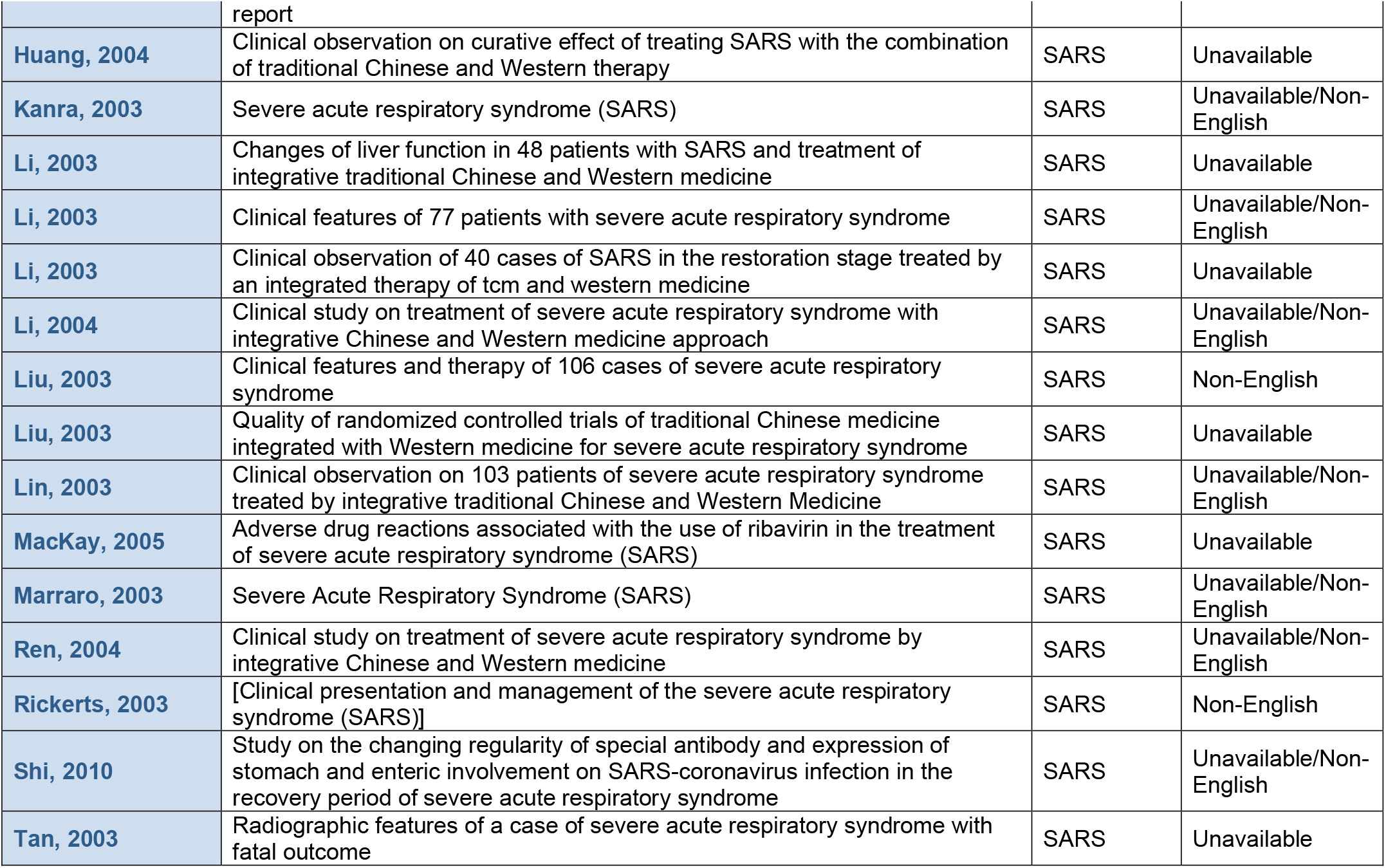
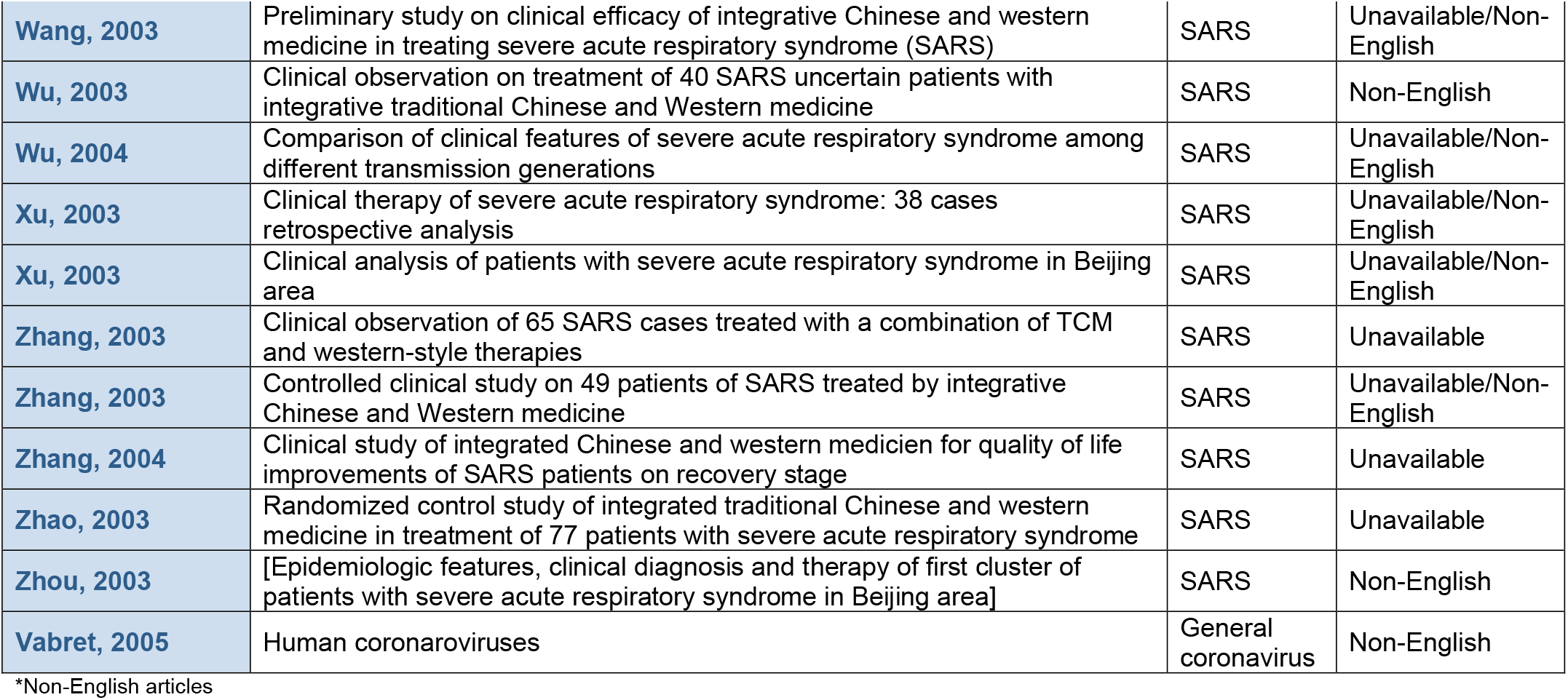

## APPENDIX 3 Detailed Table of Study and Patient Characteristics

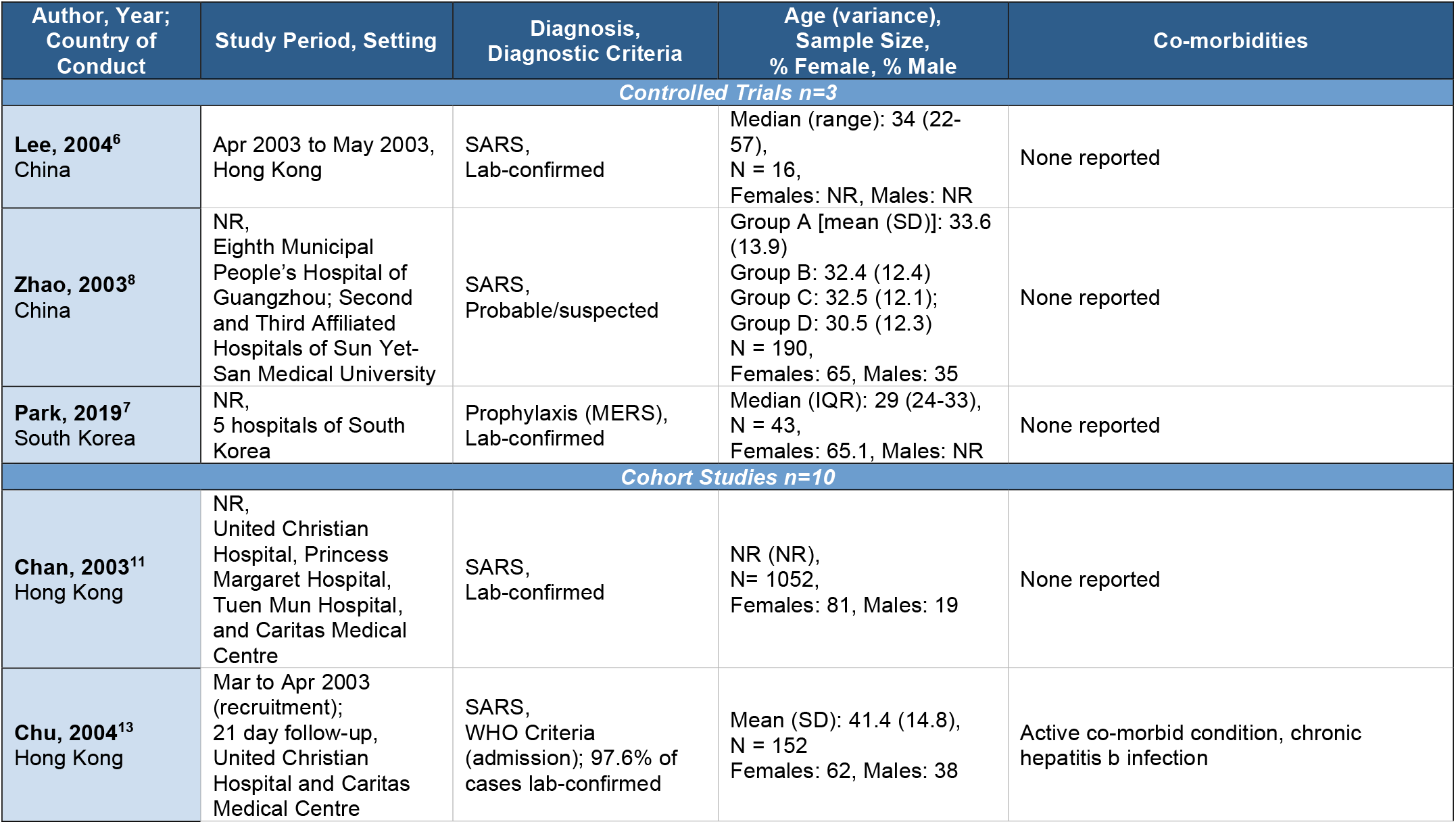
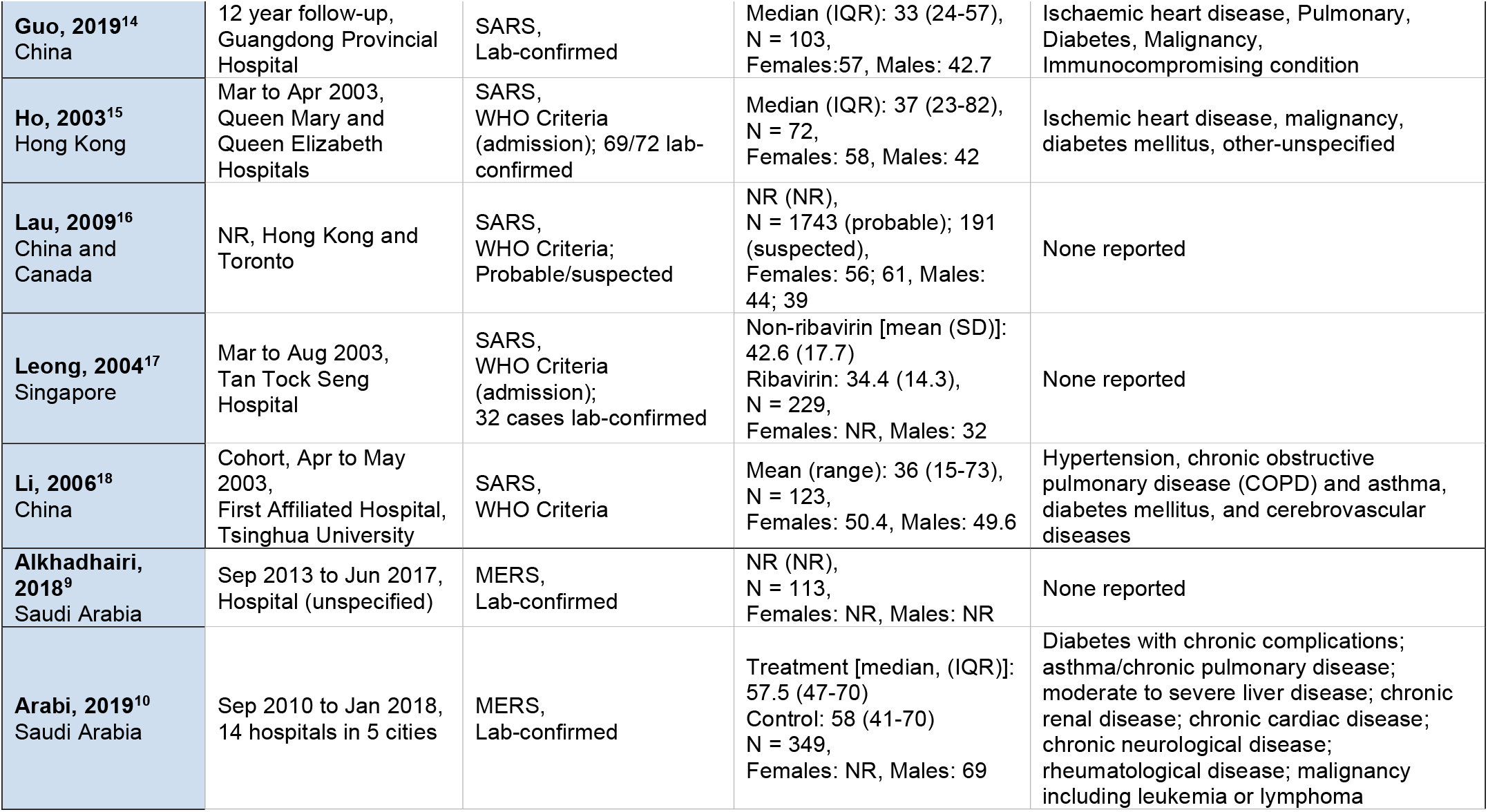
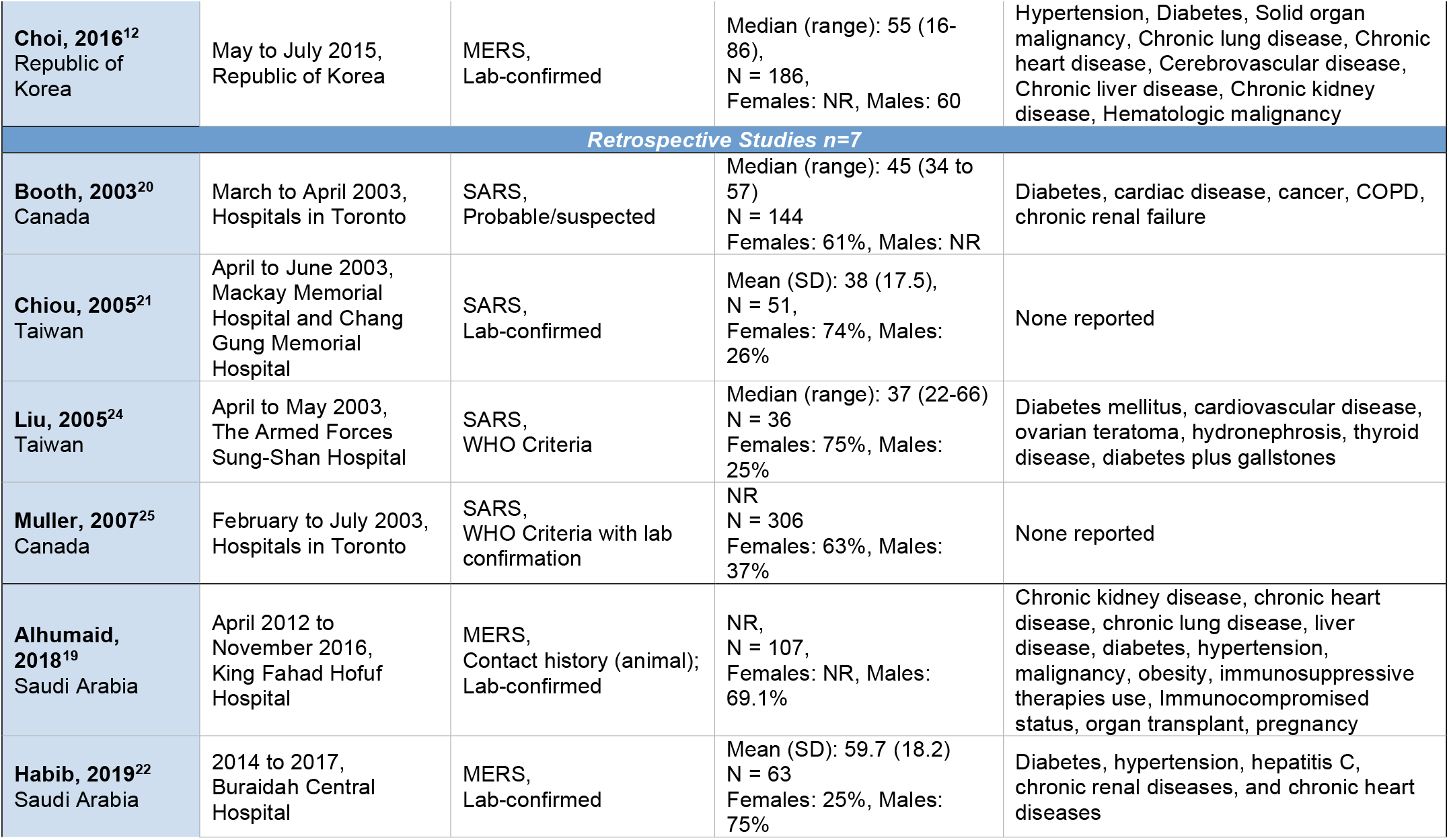
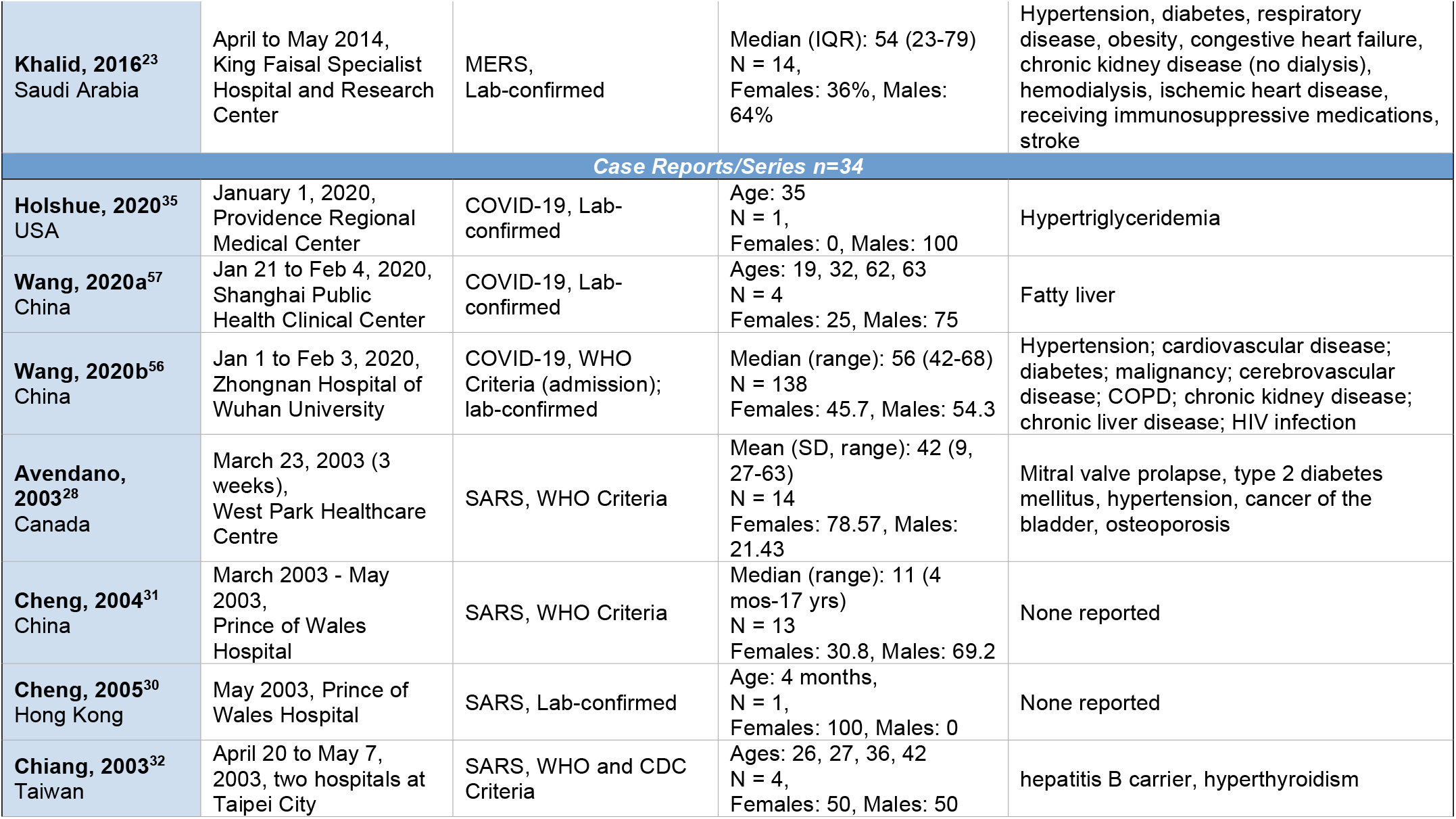
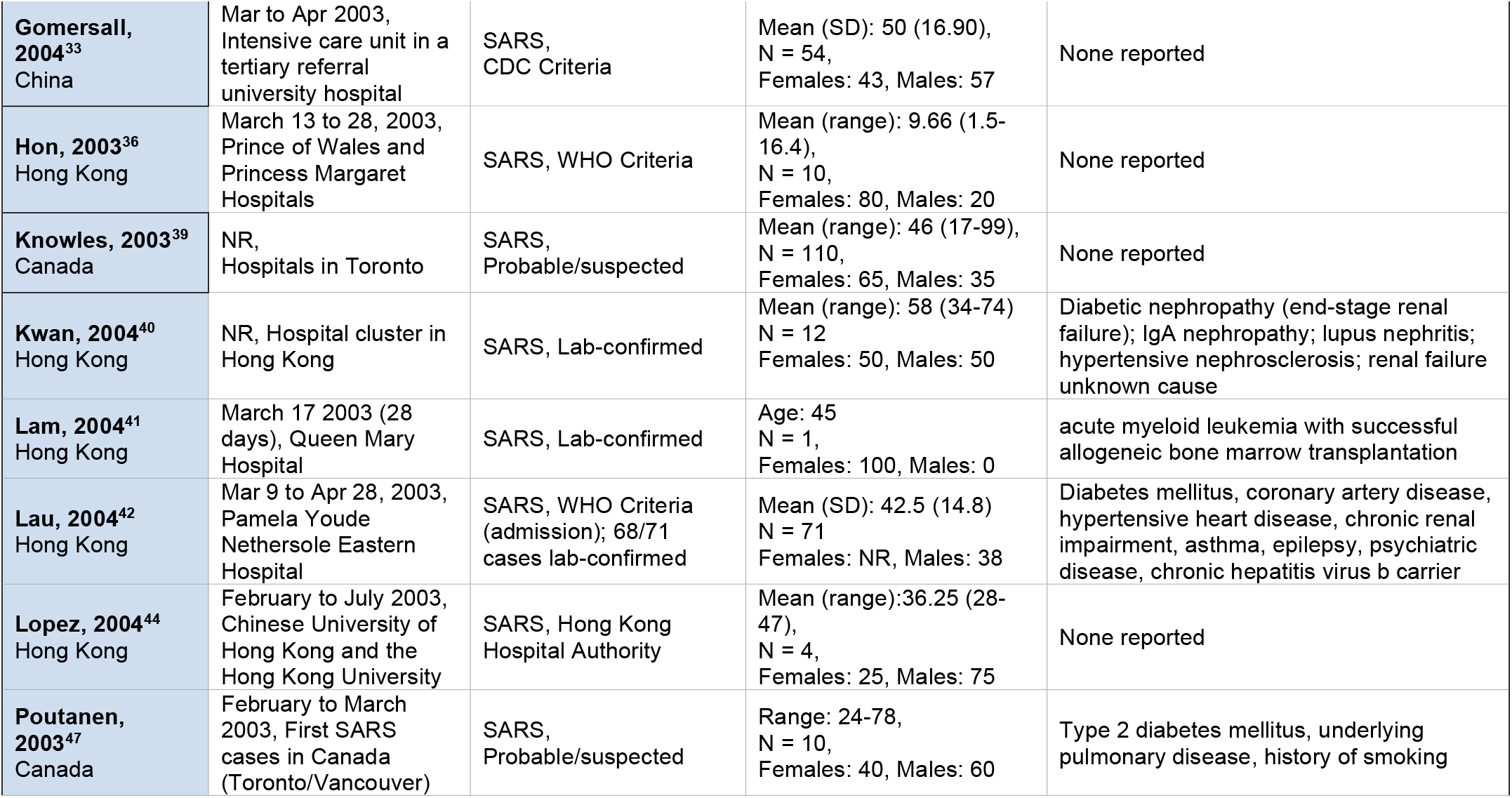
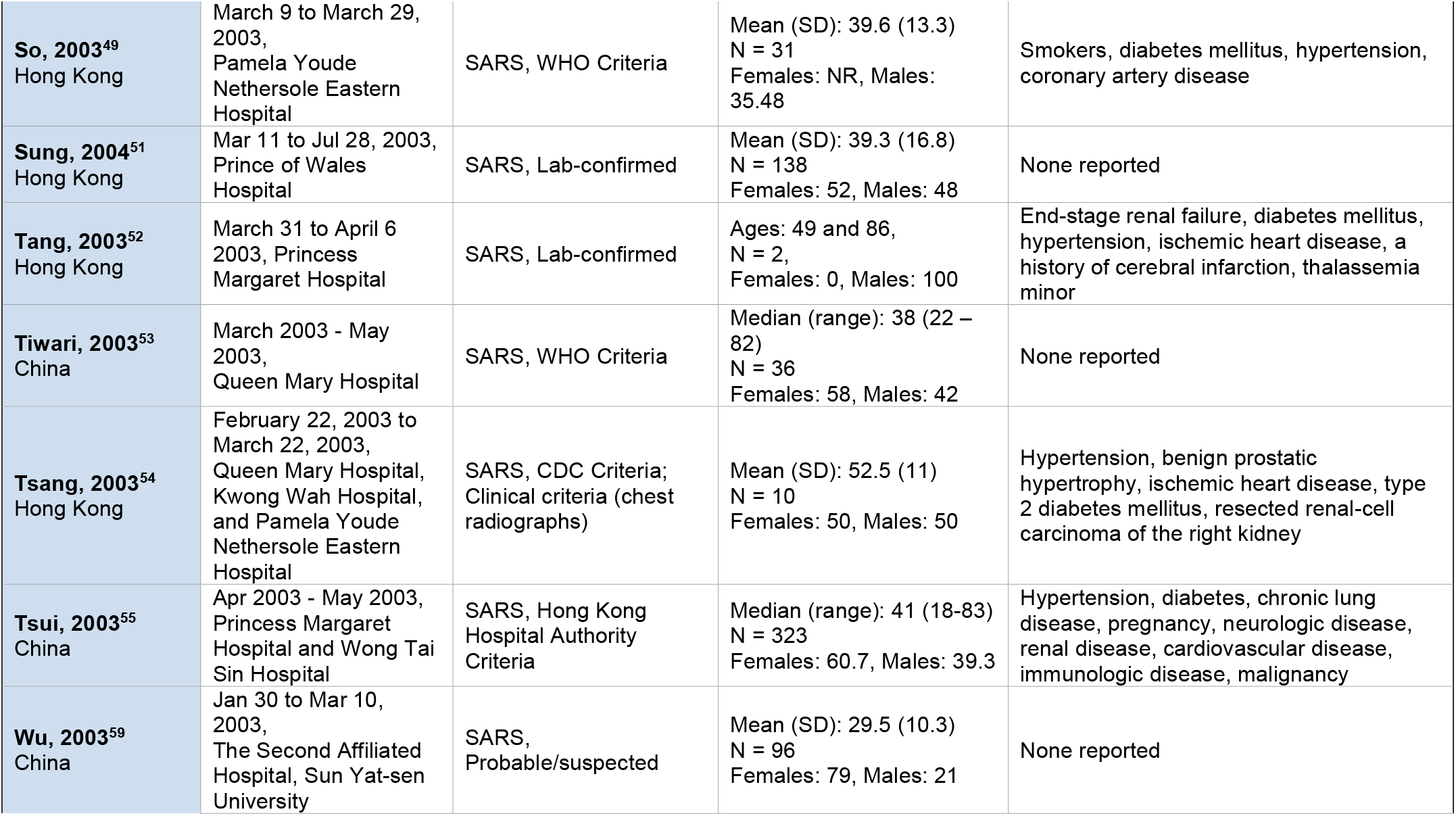
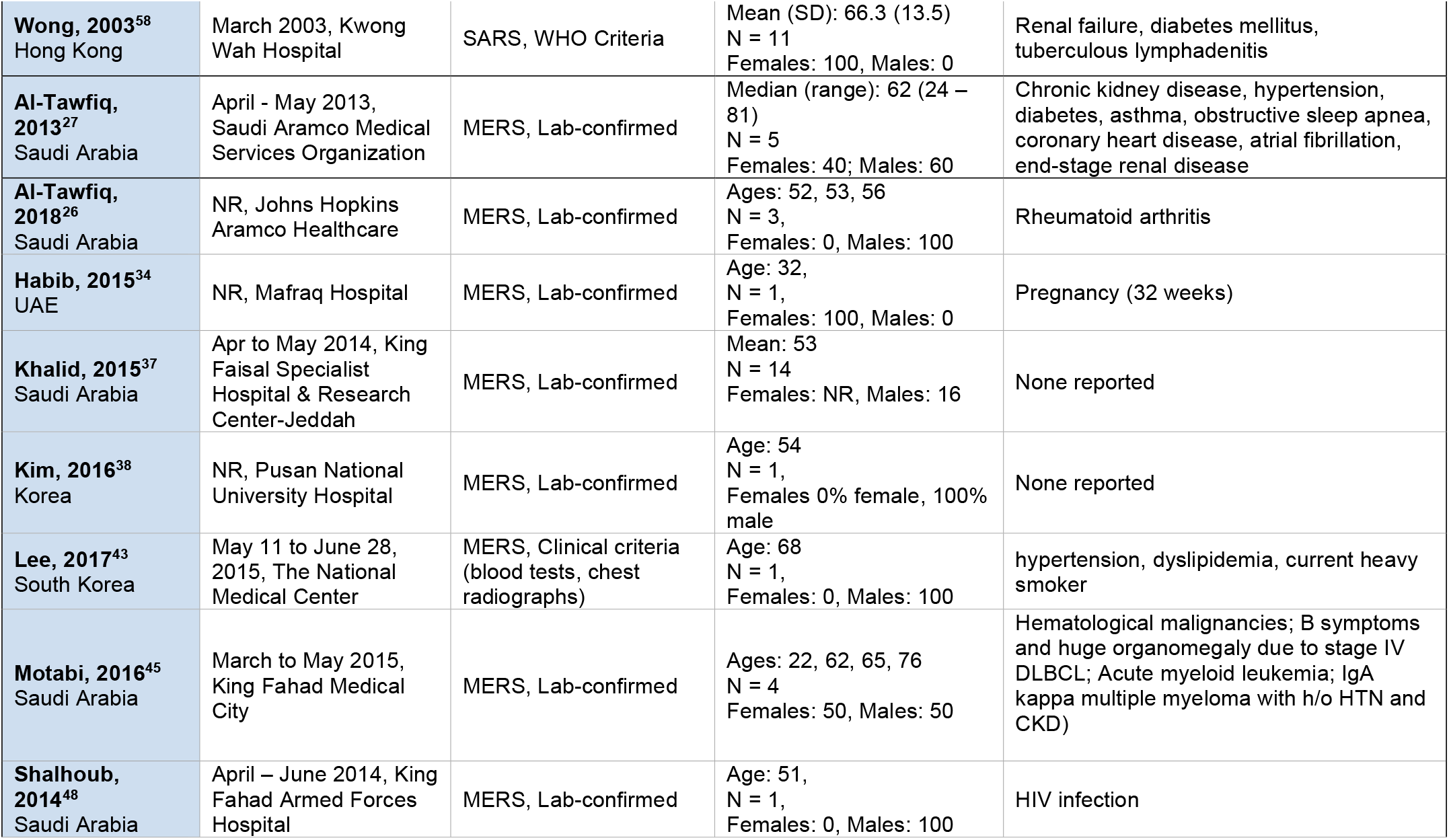
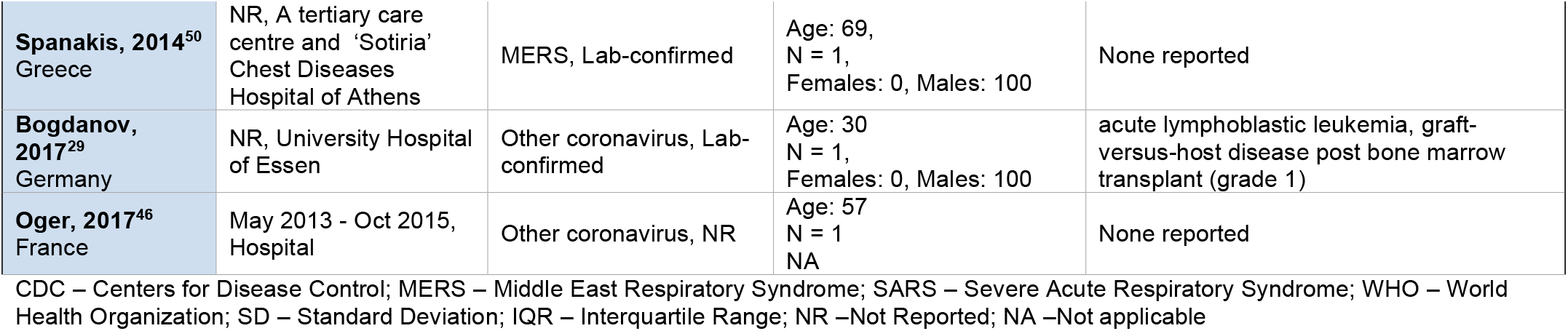

## APPENDIX 4 Detailed Table of Interventions and Outcomes

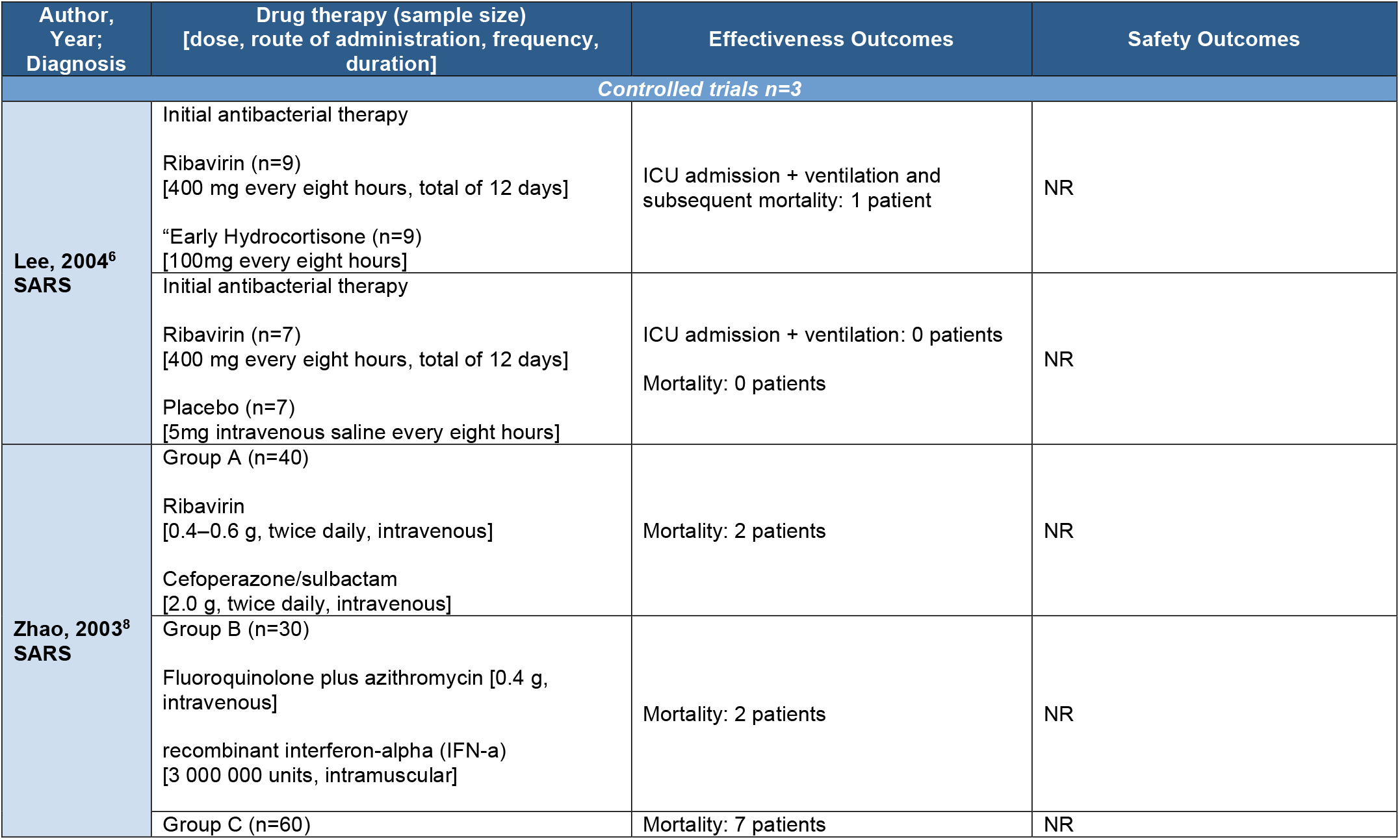
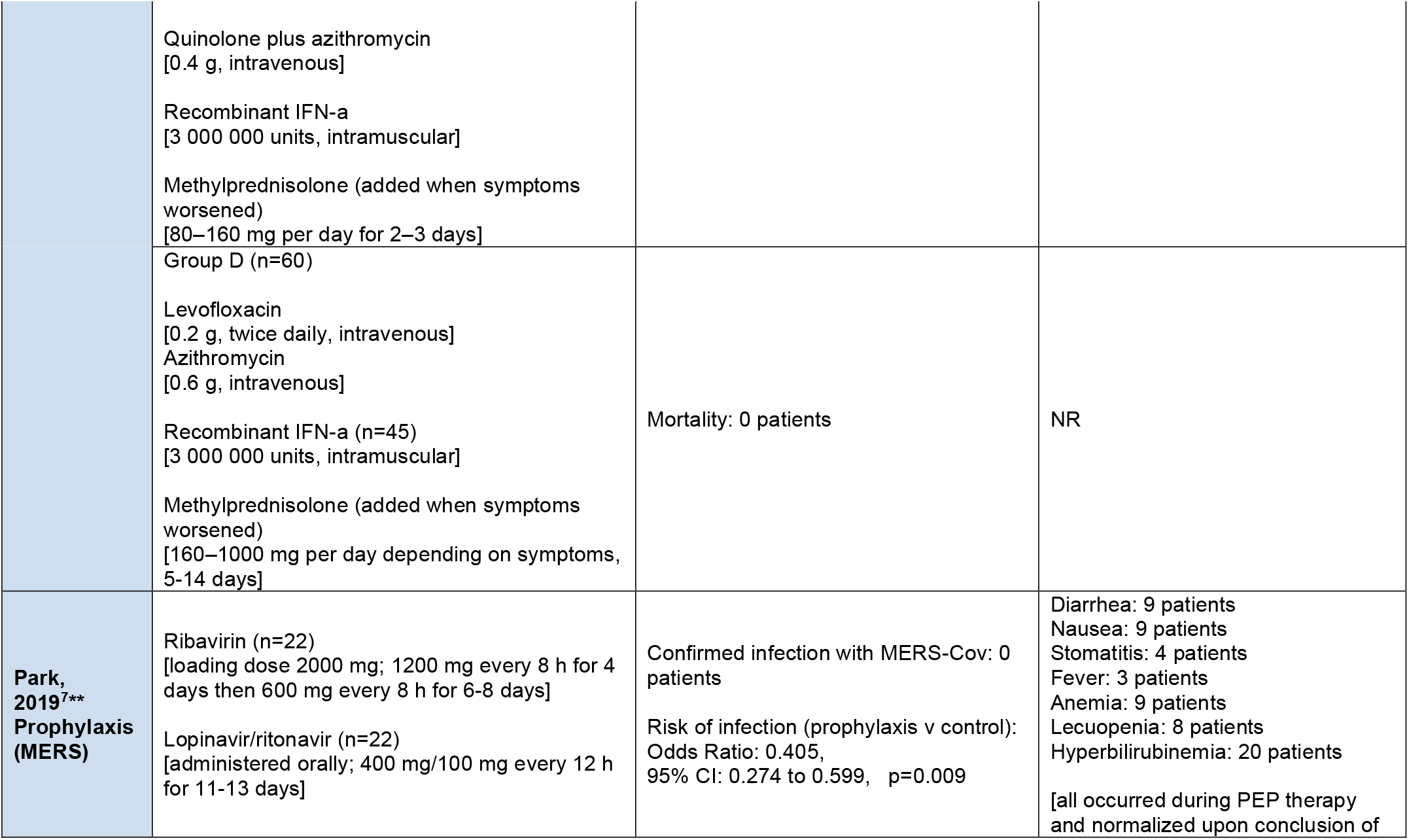
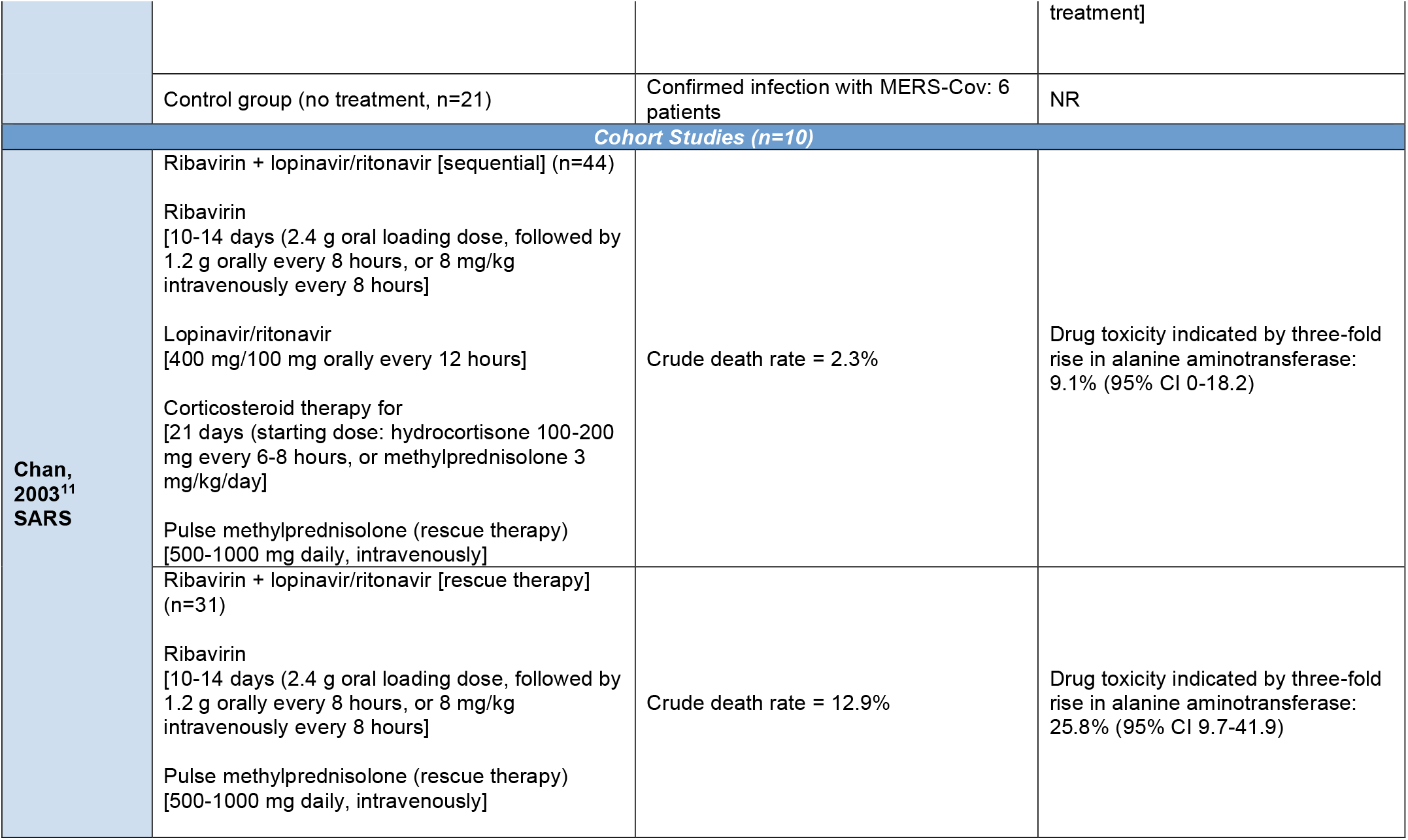
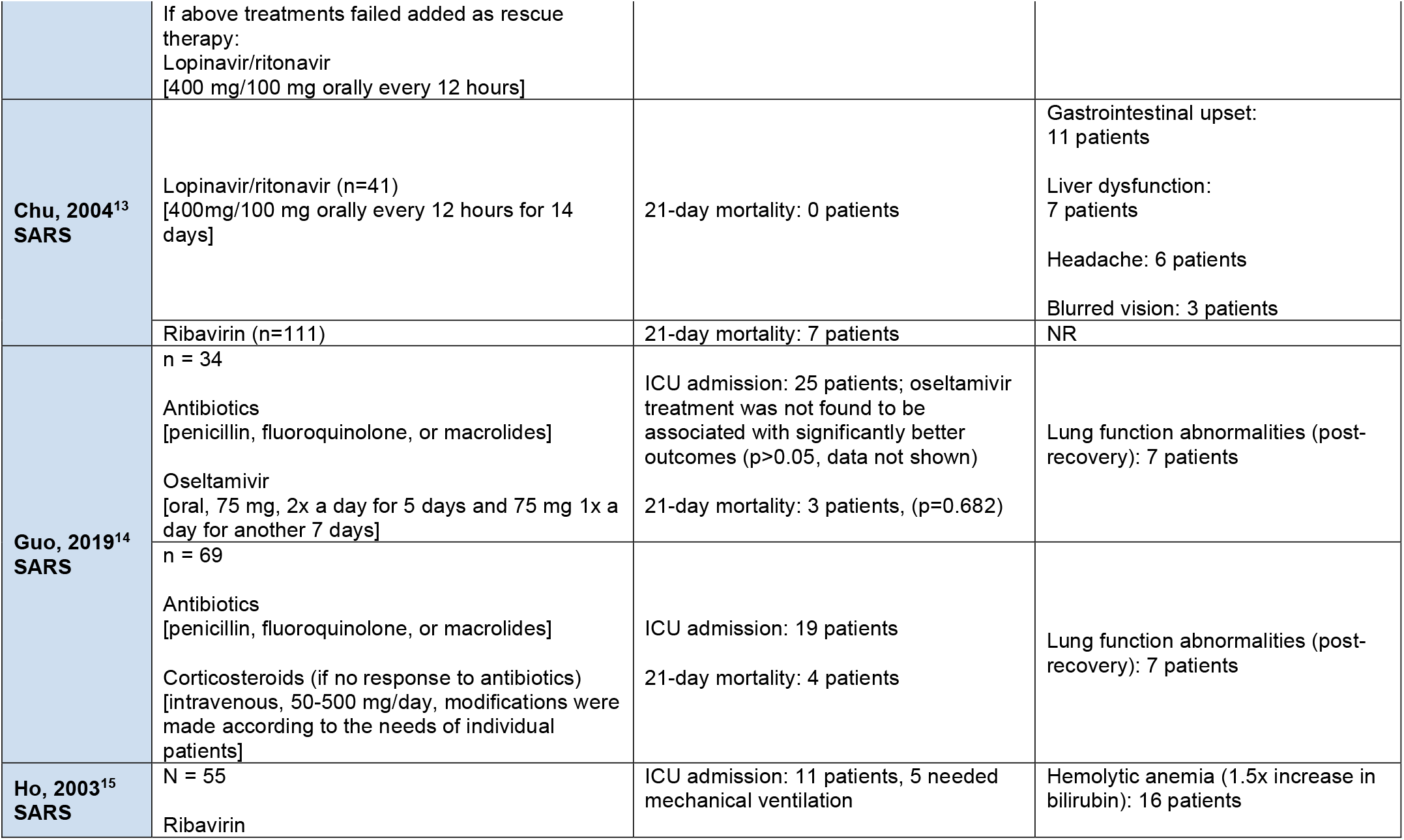
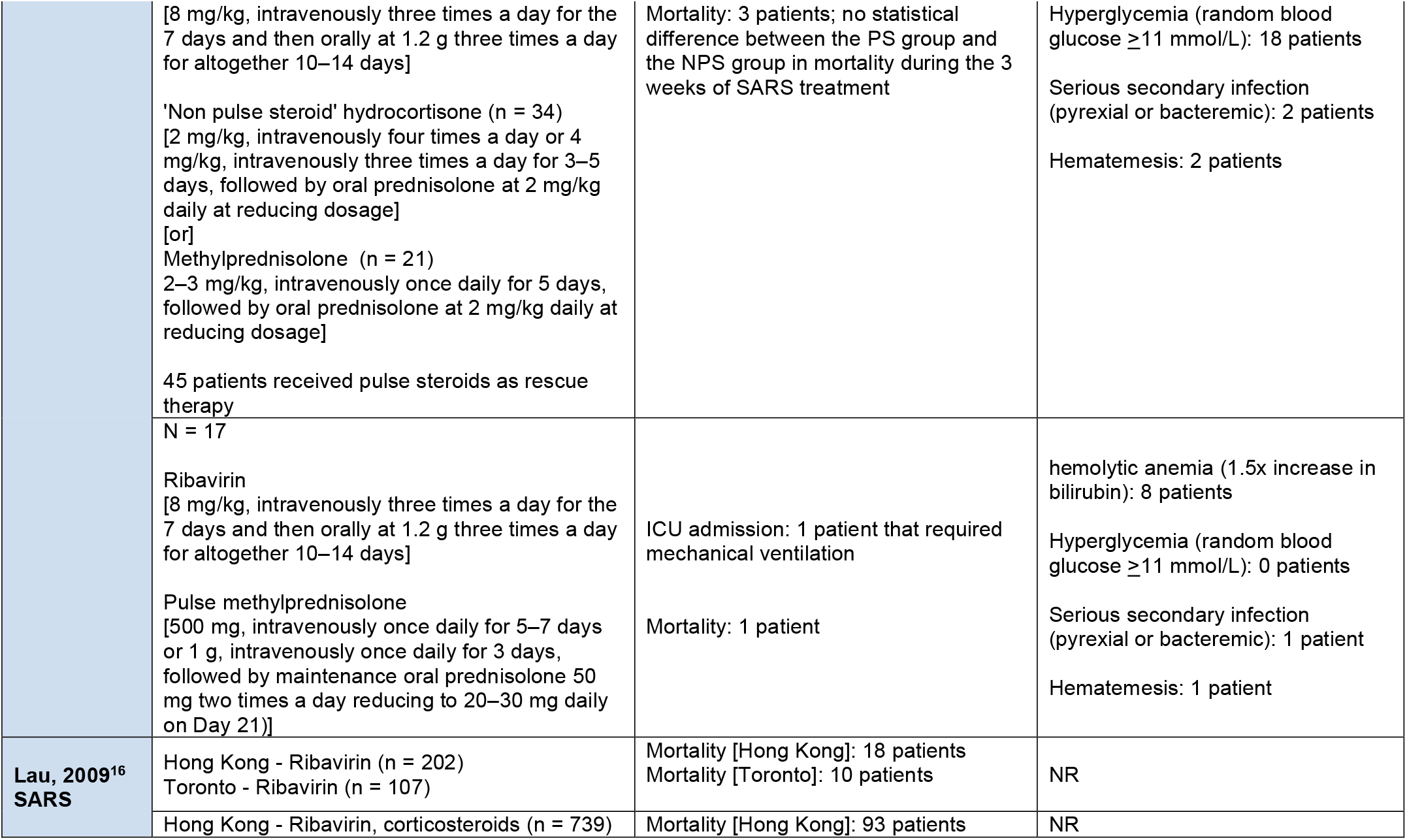
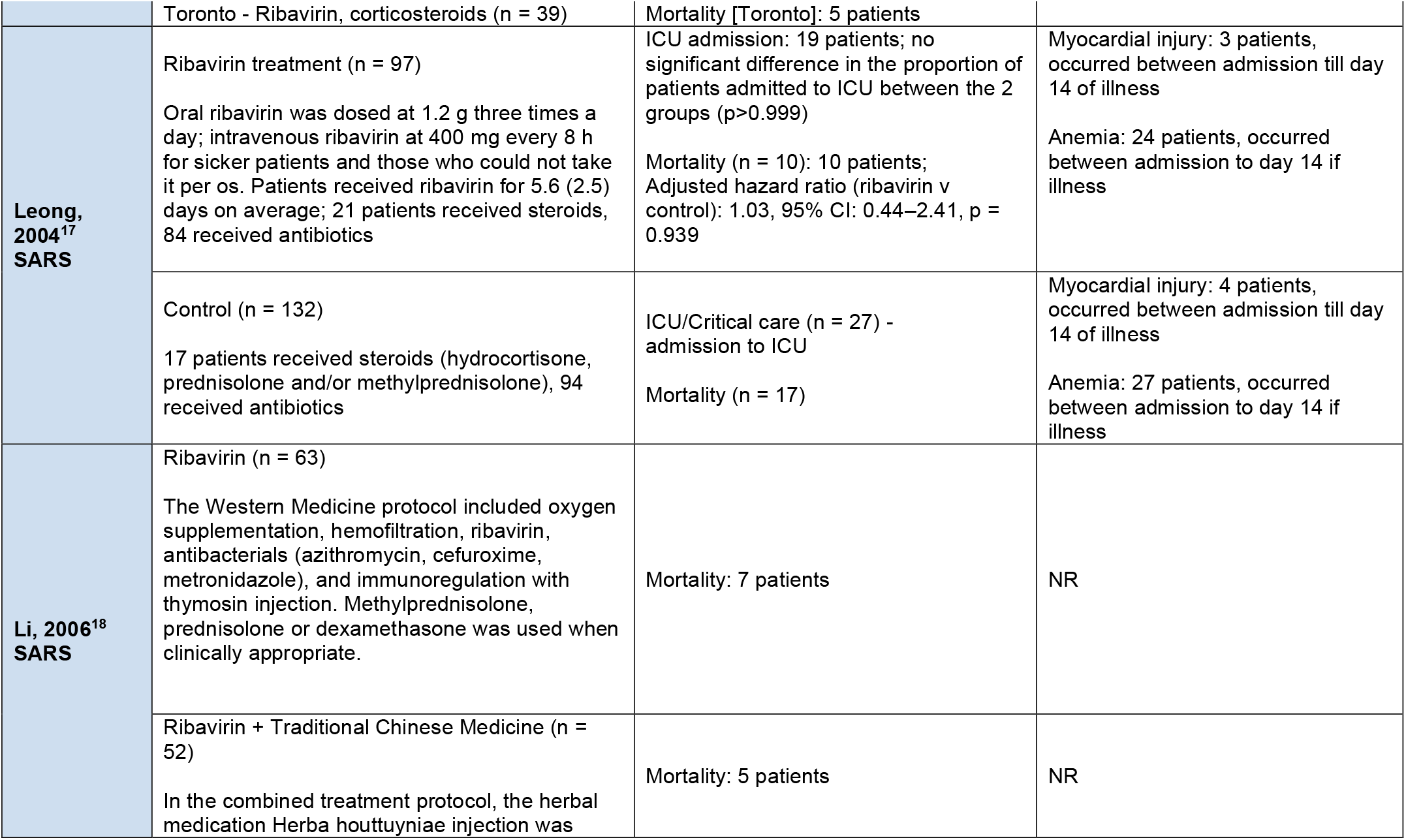
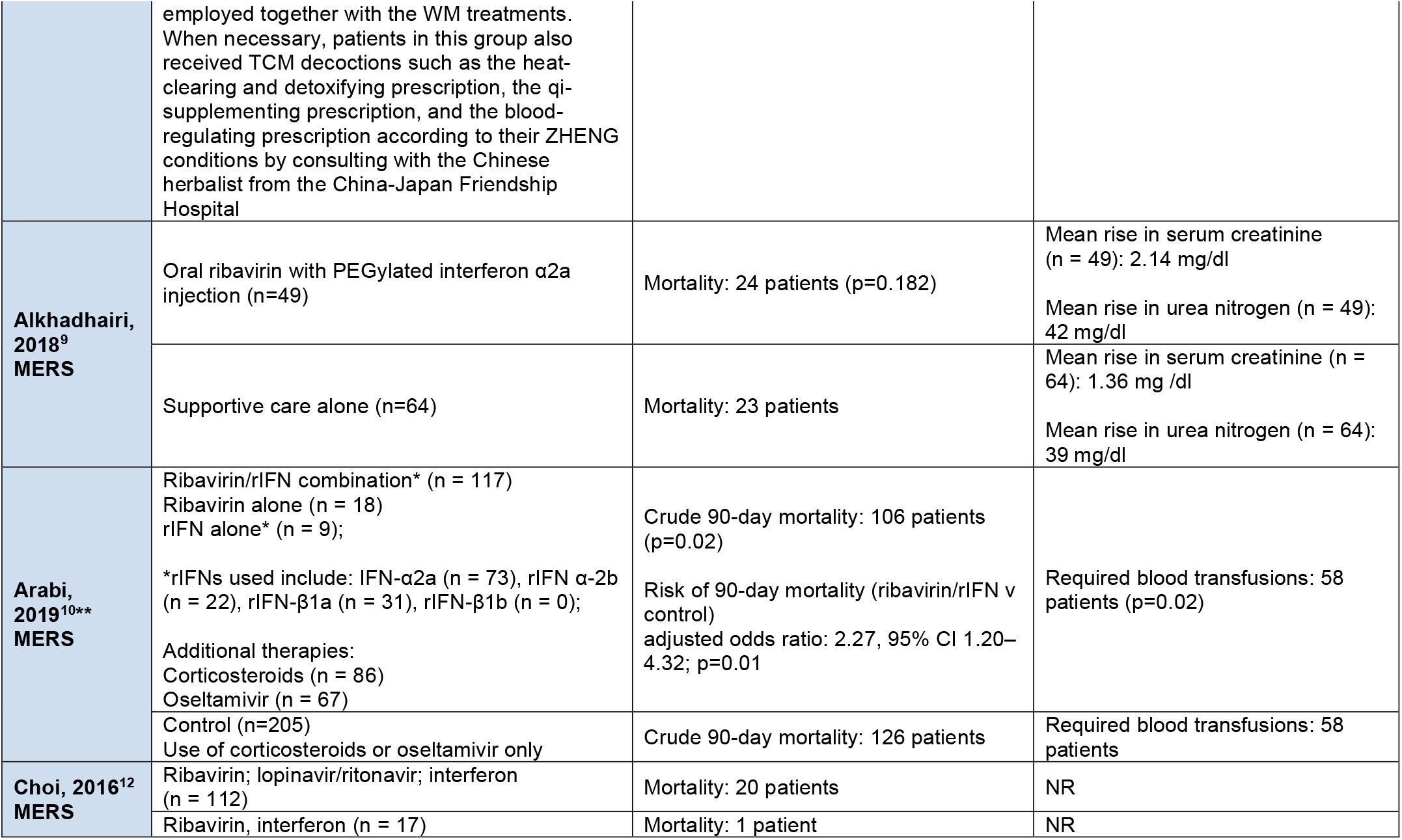
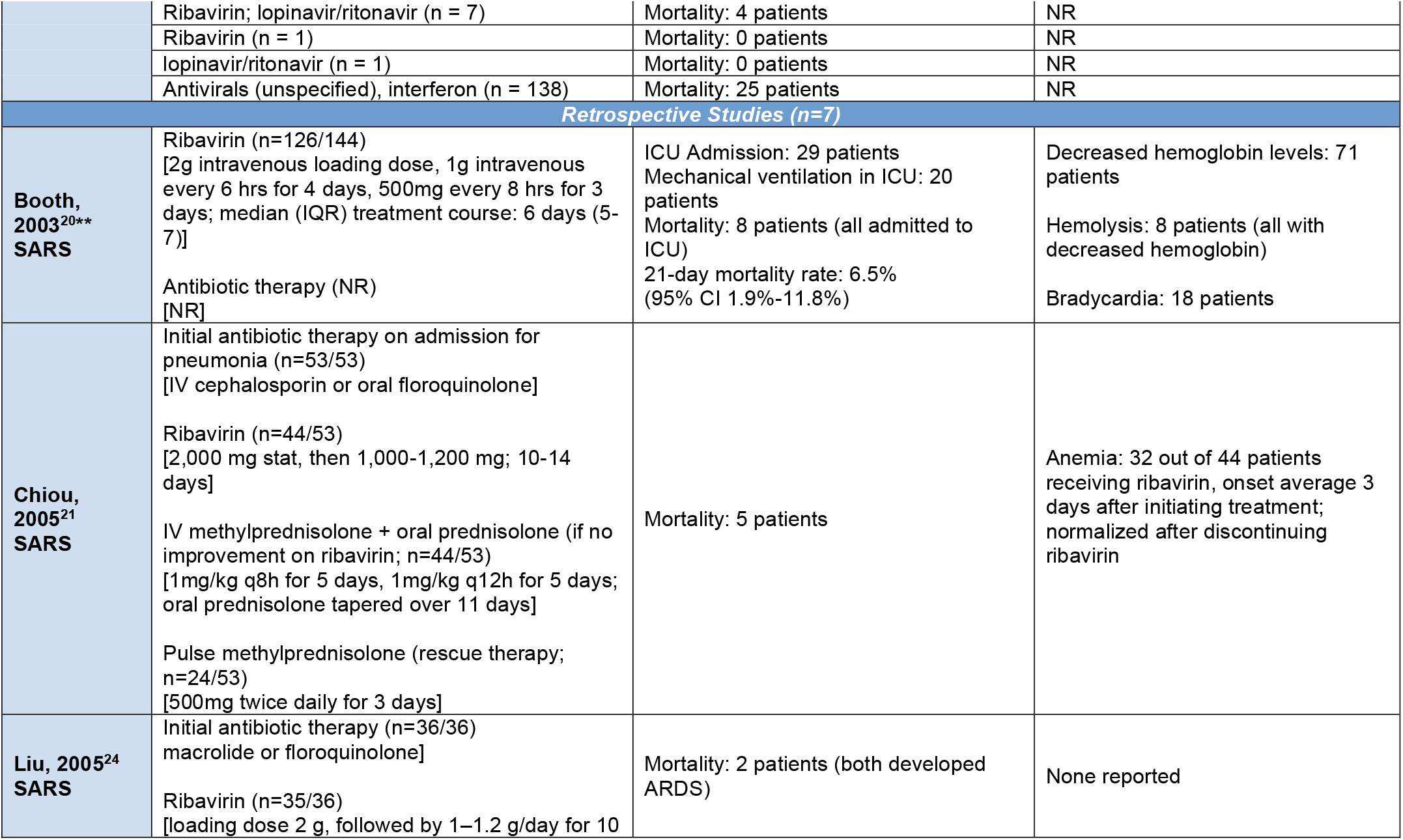
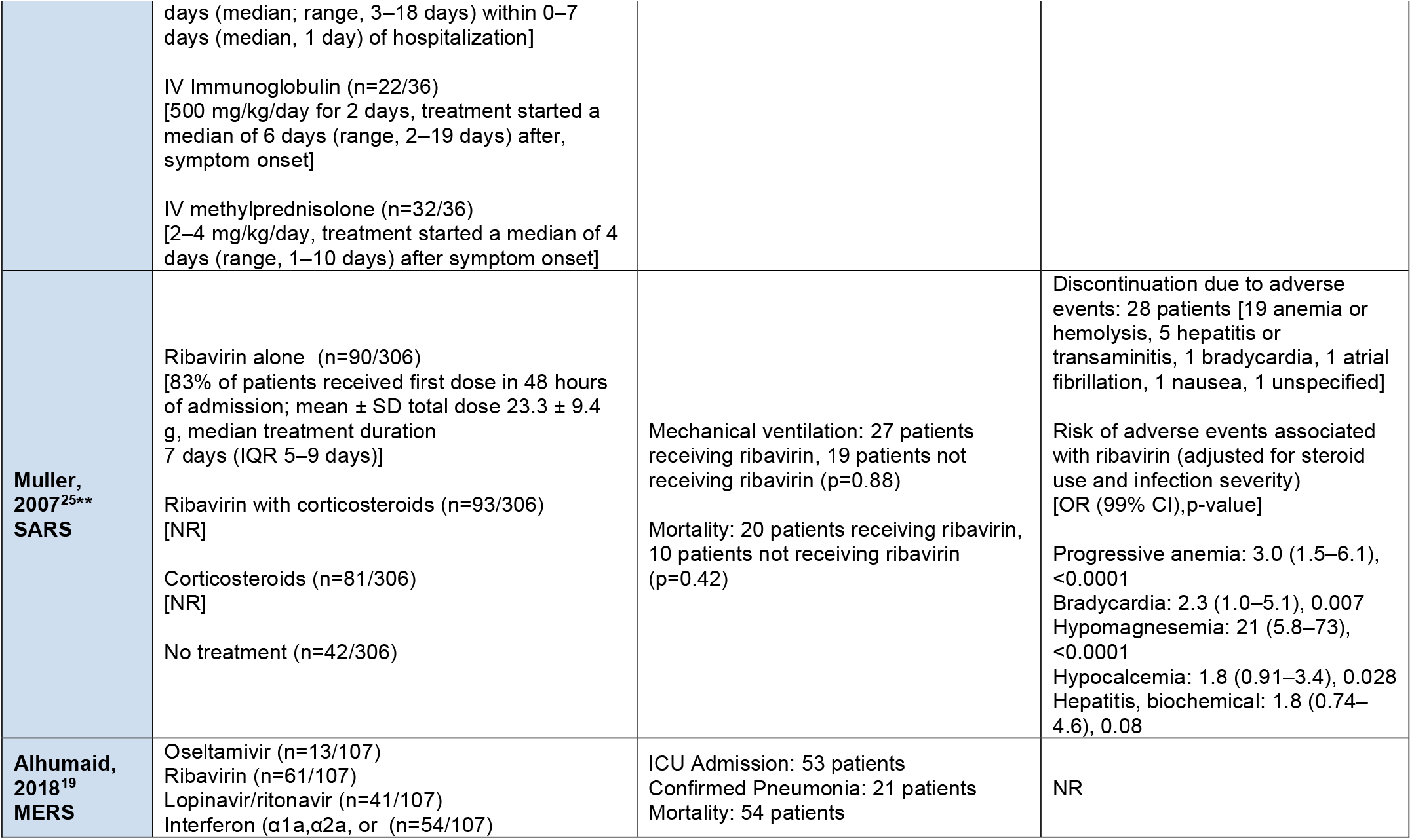
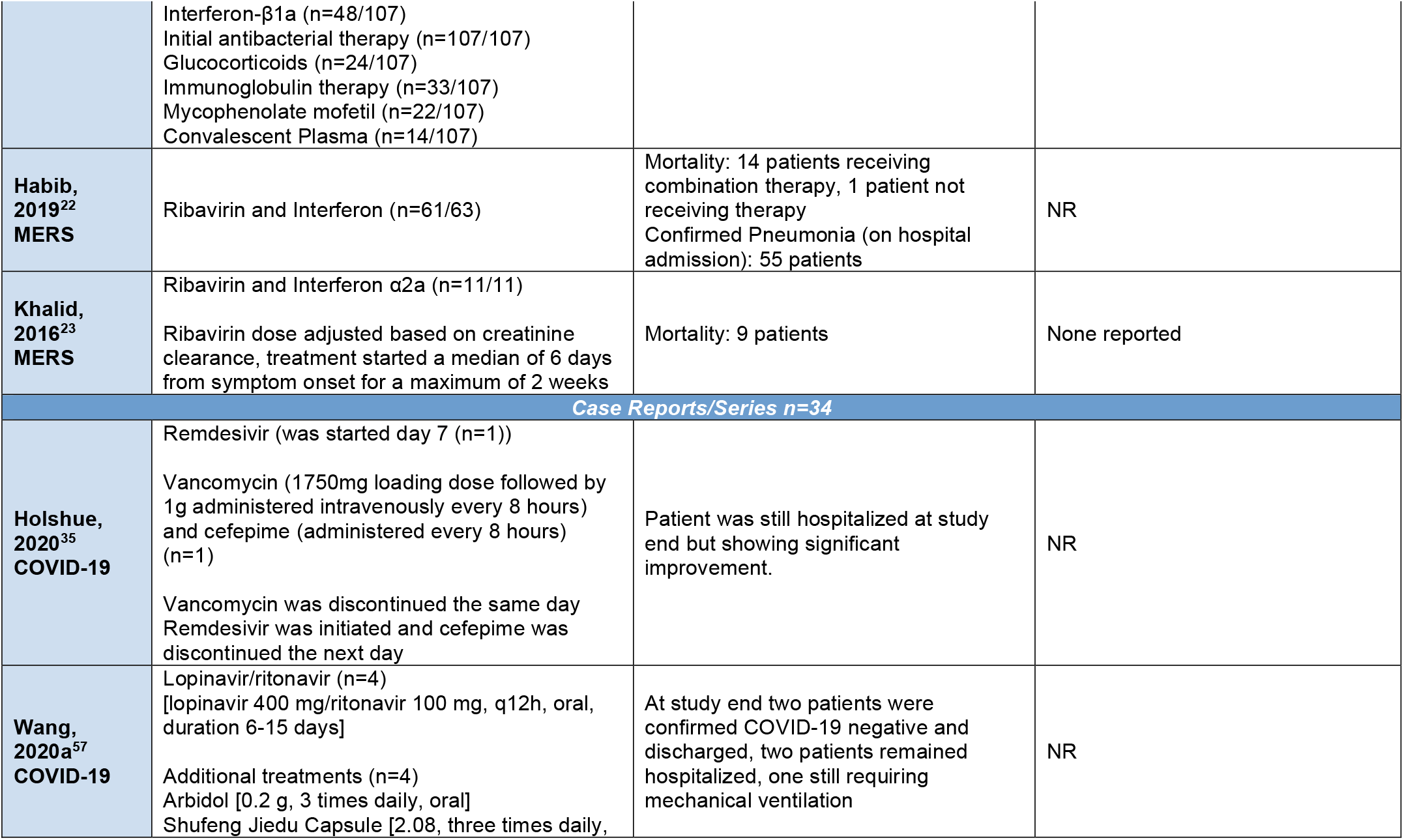
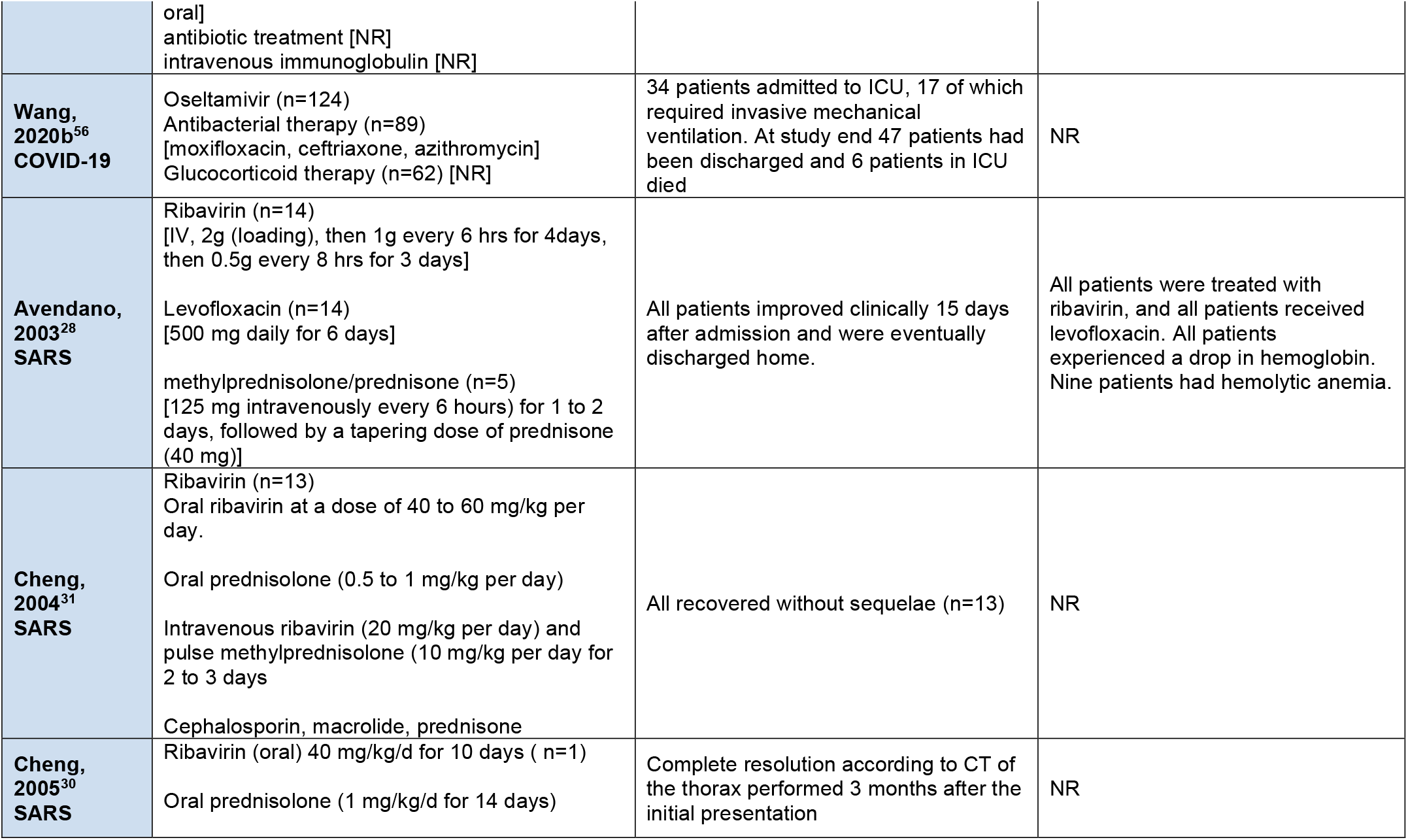
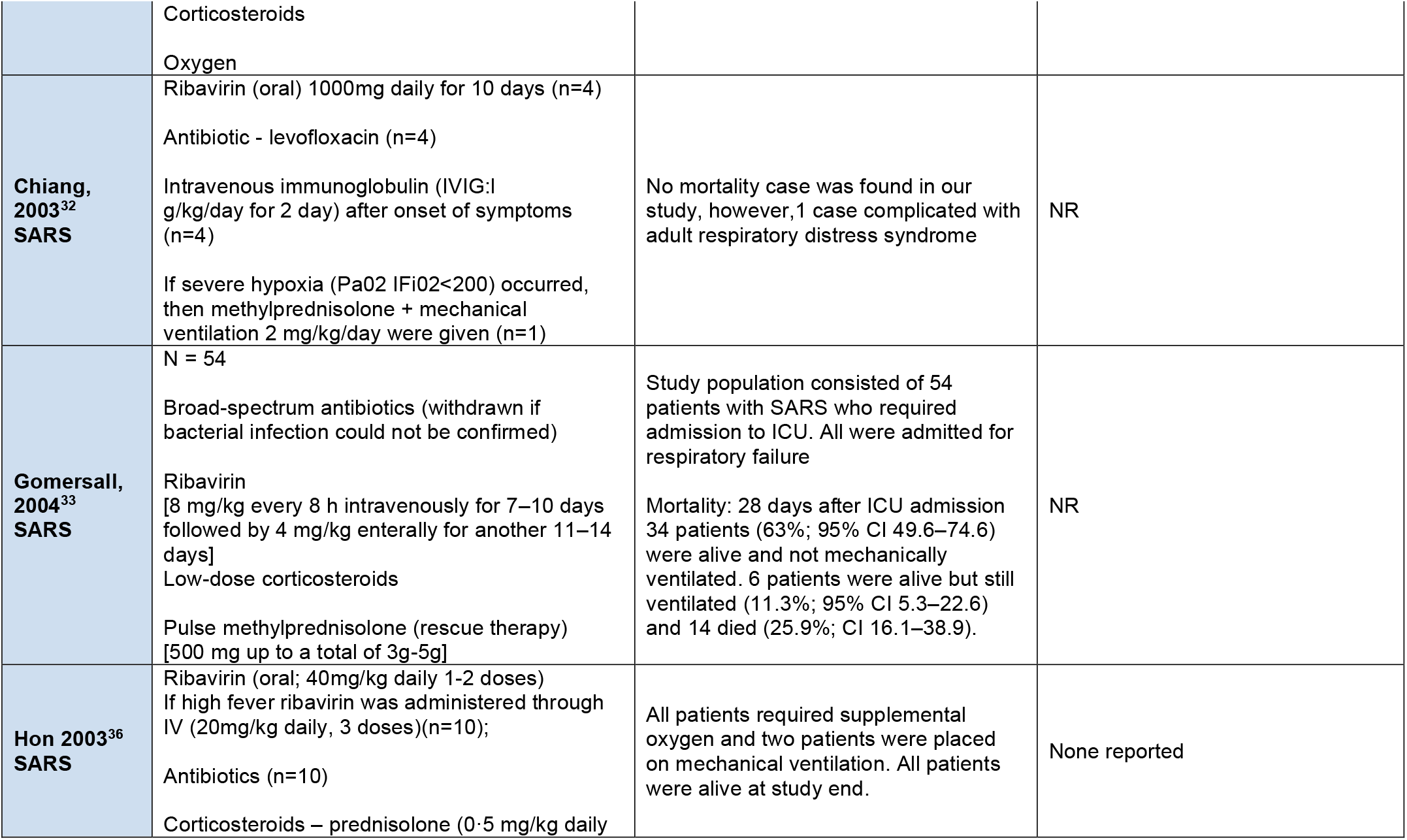
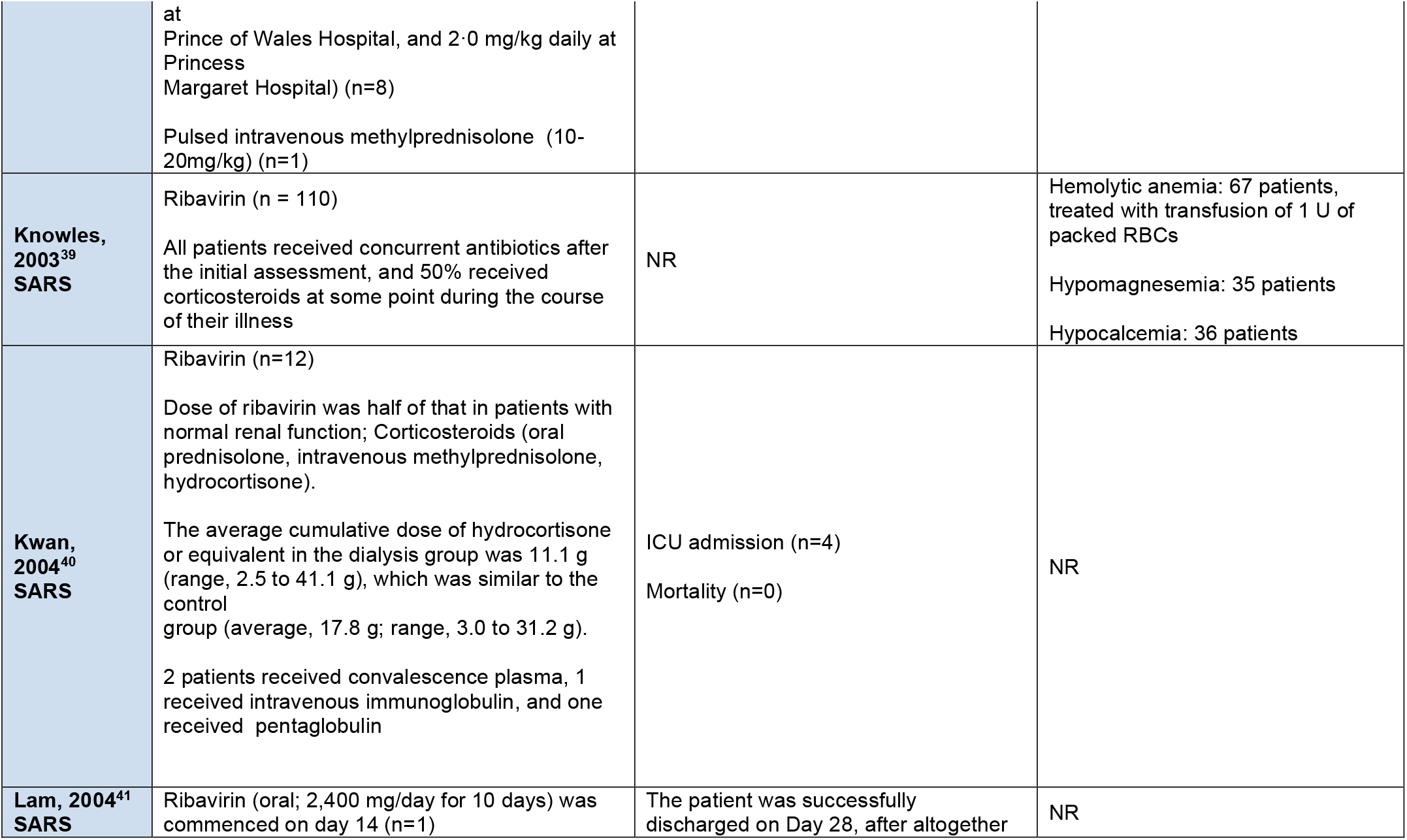
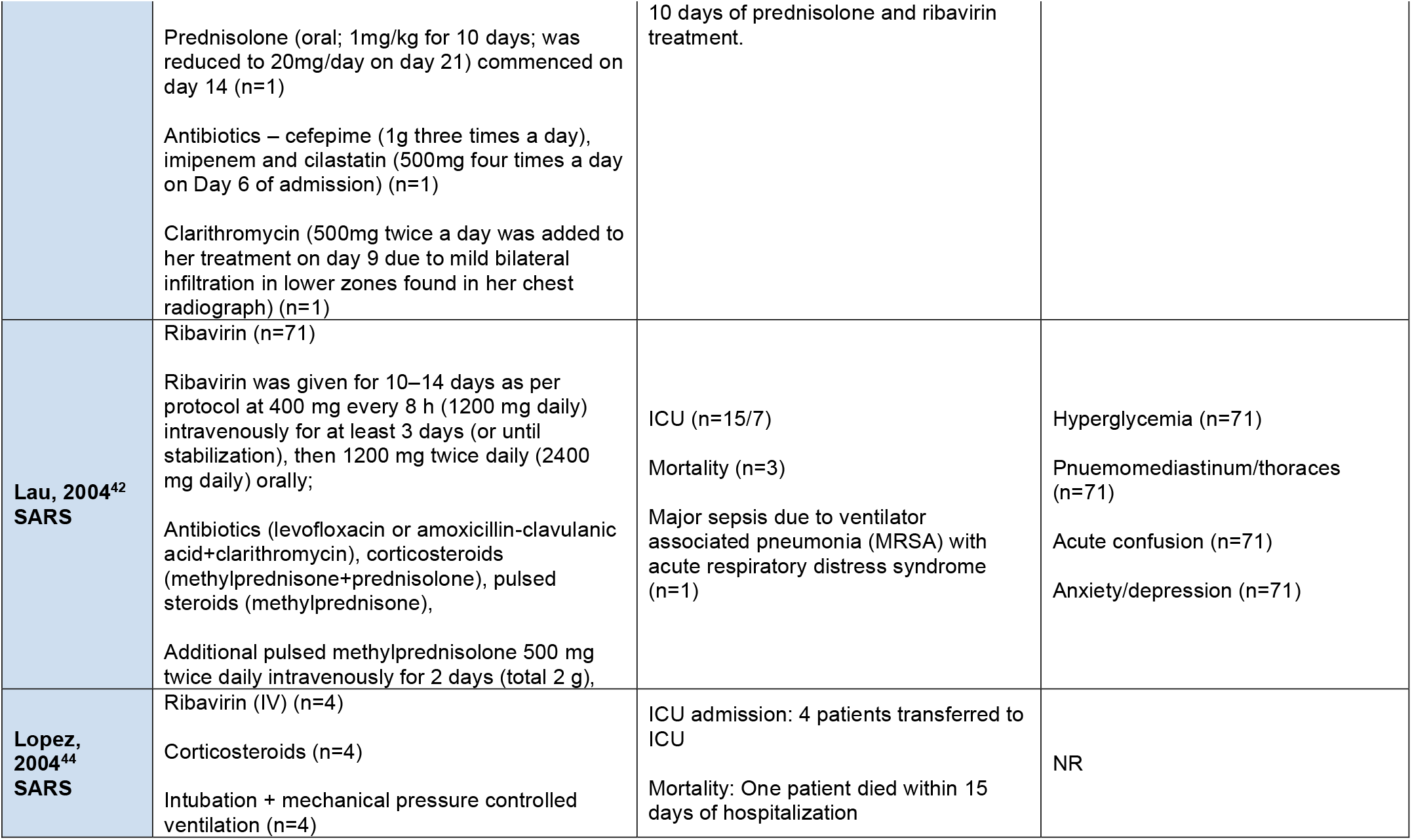
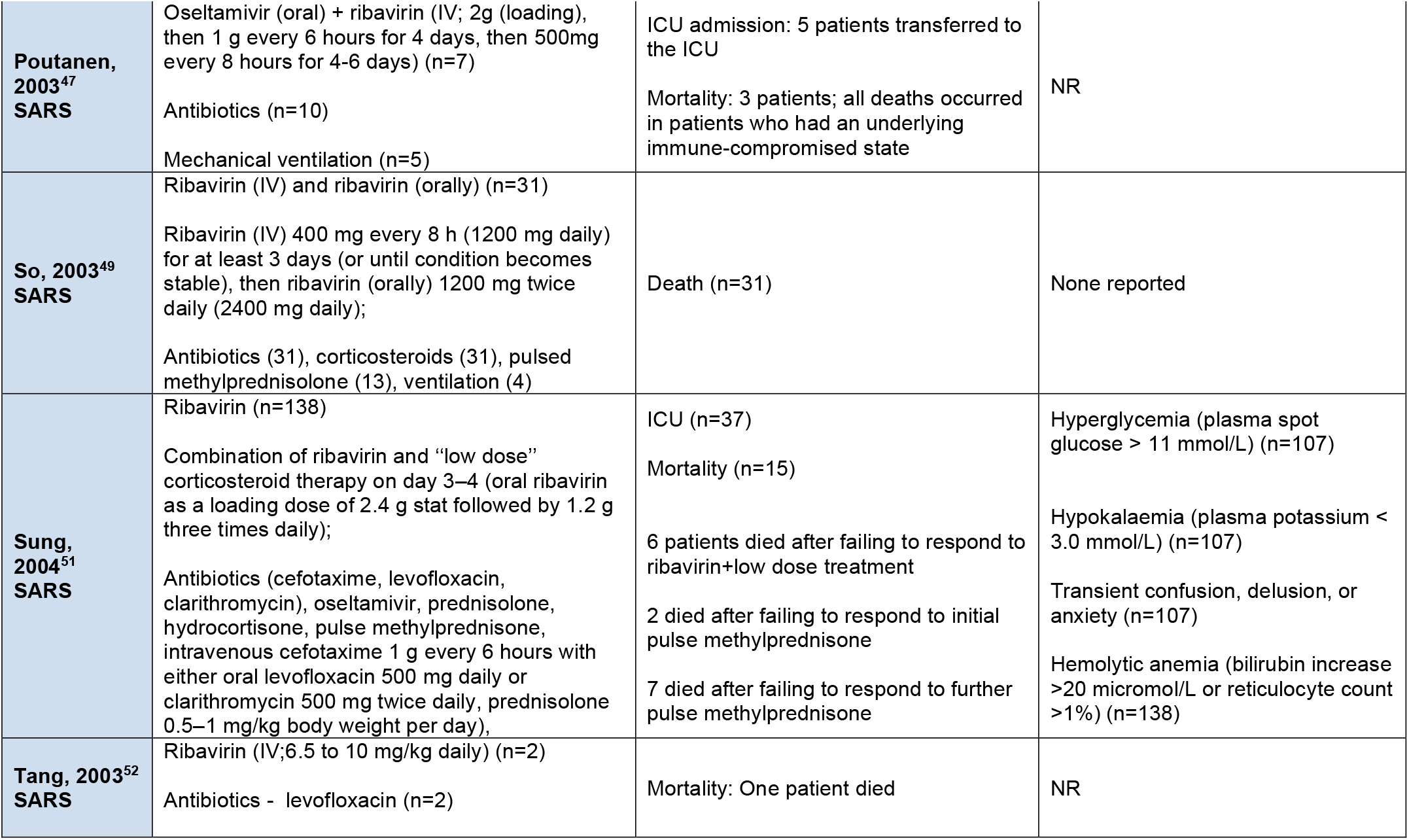
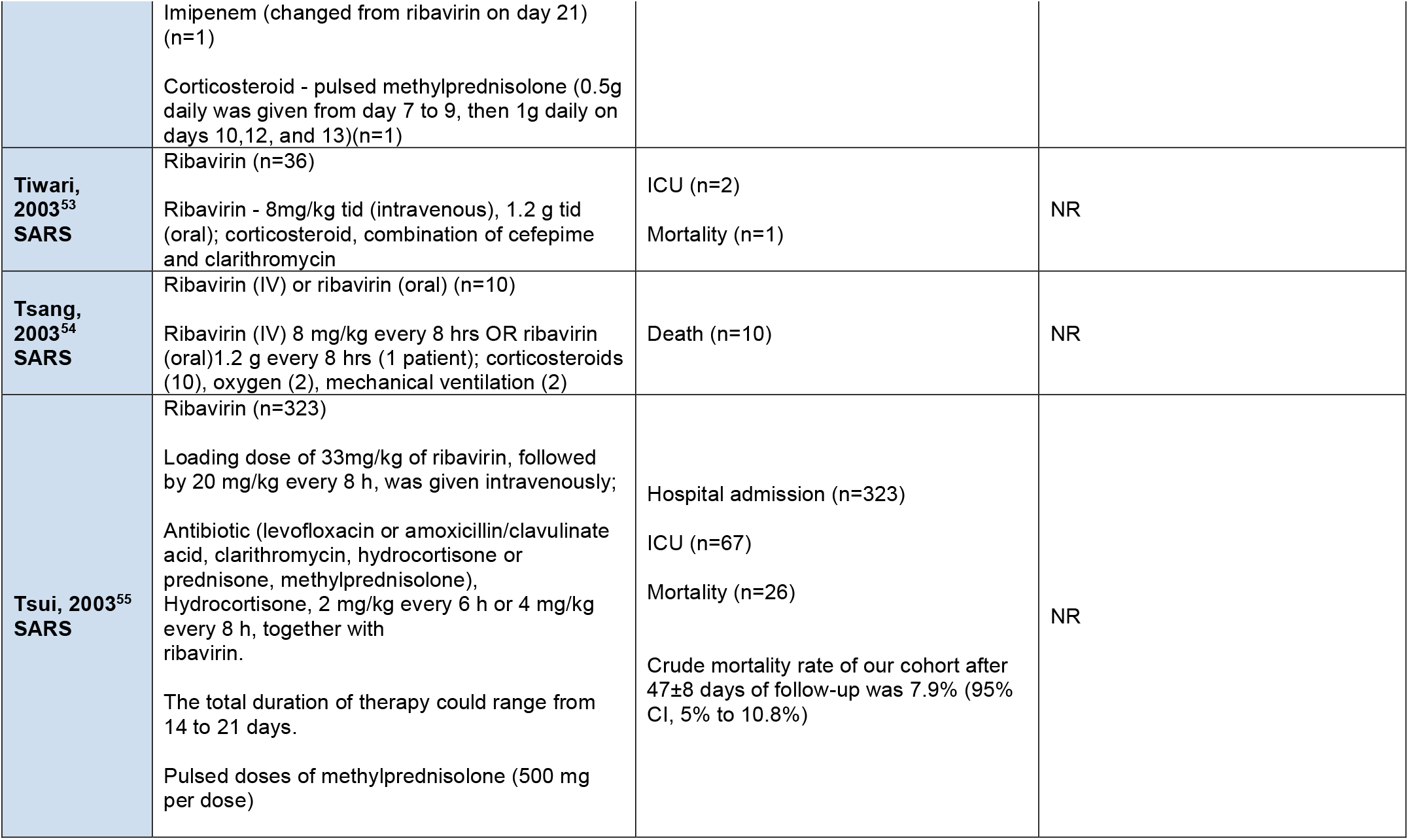
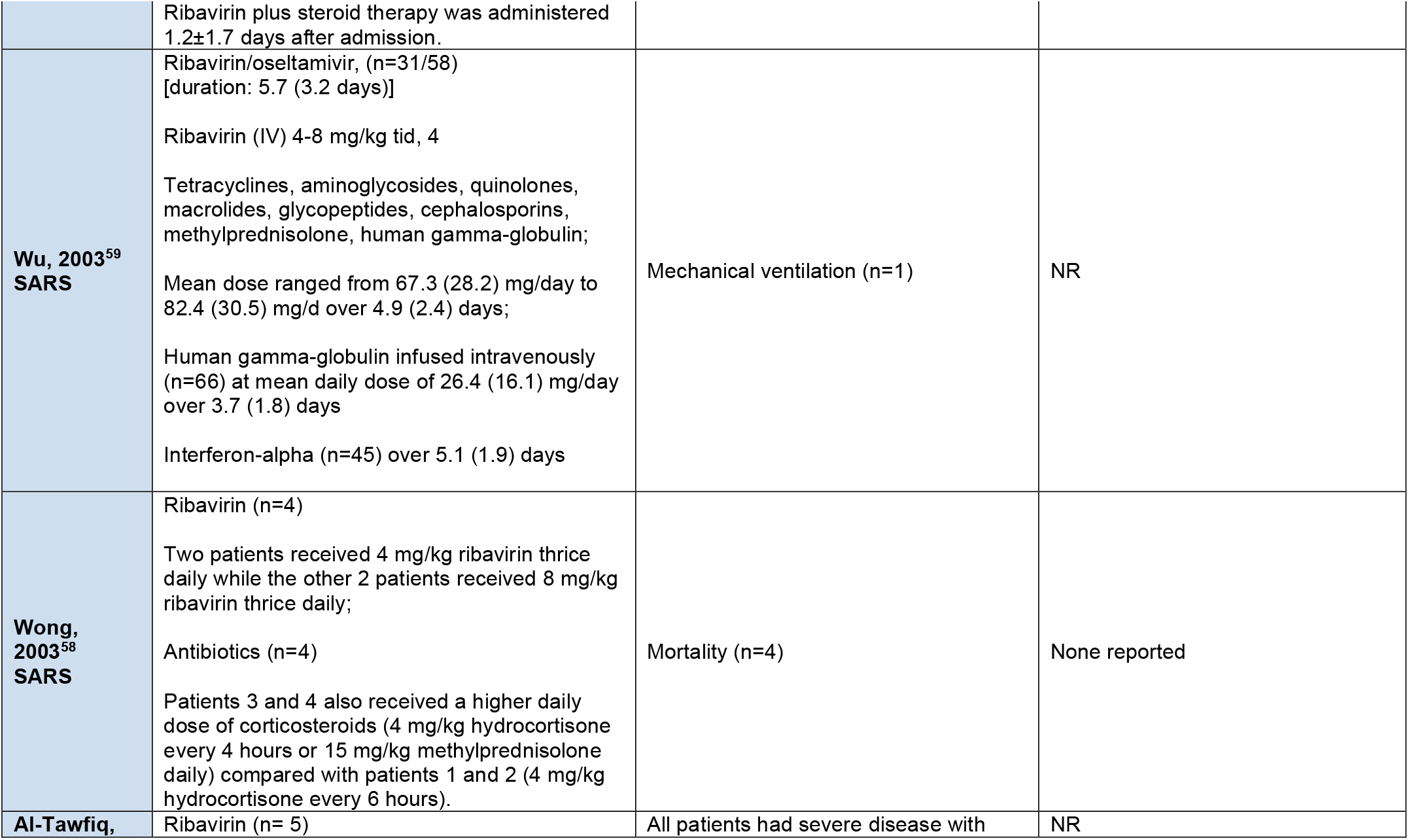
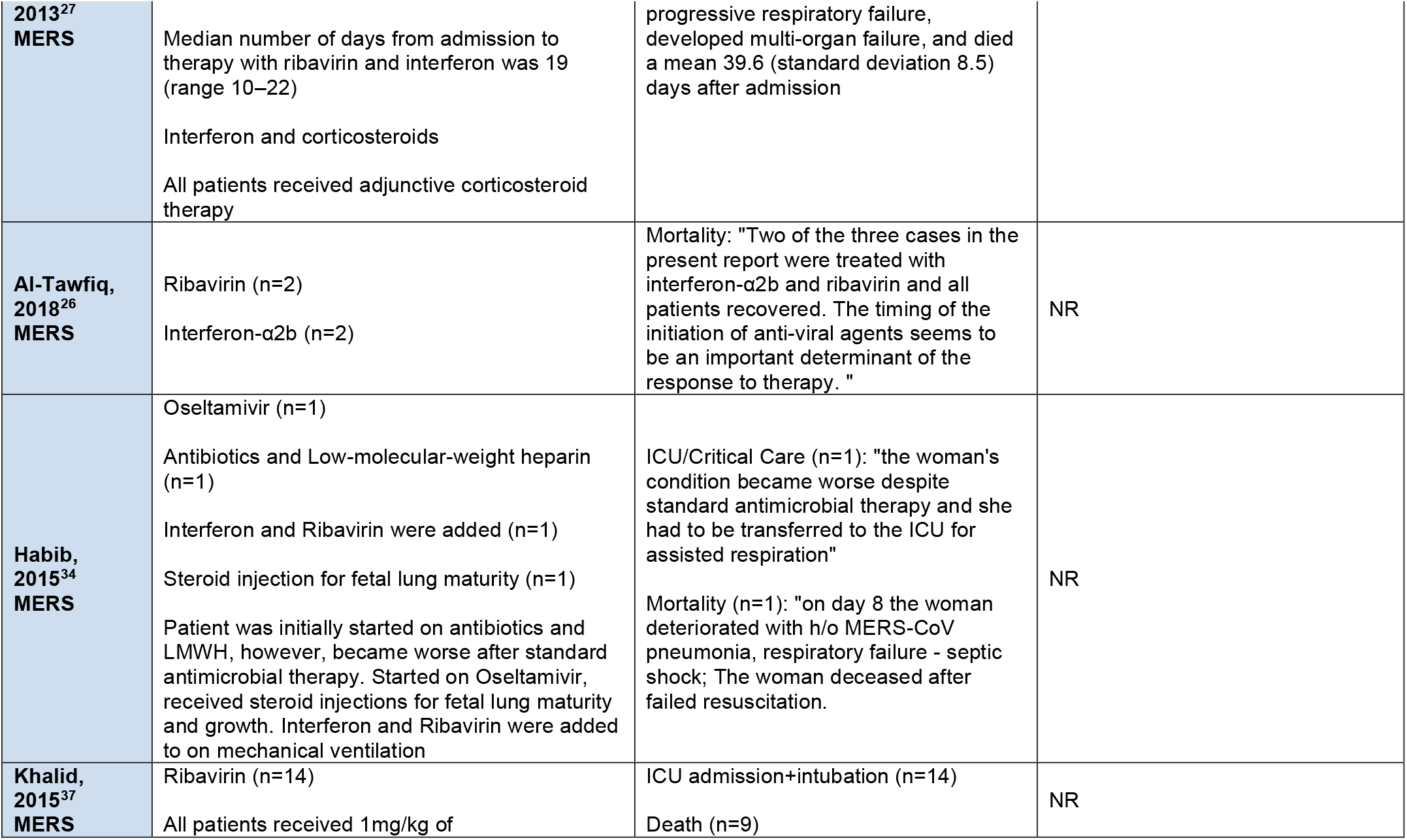
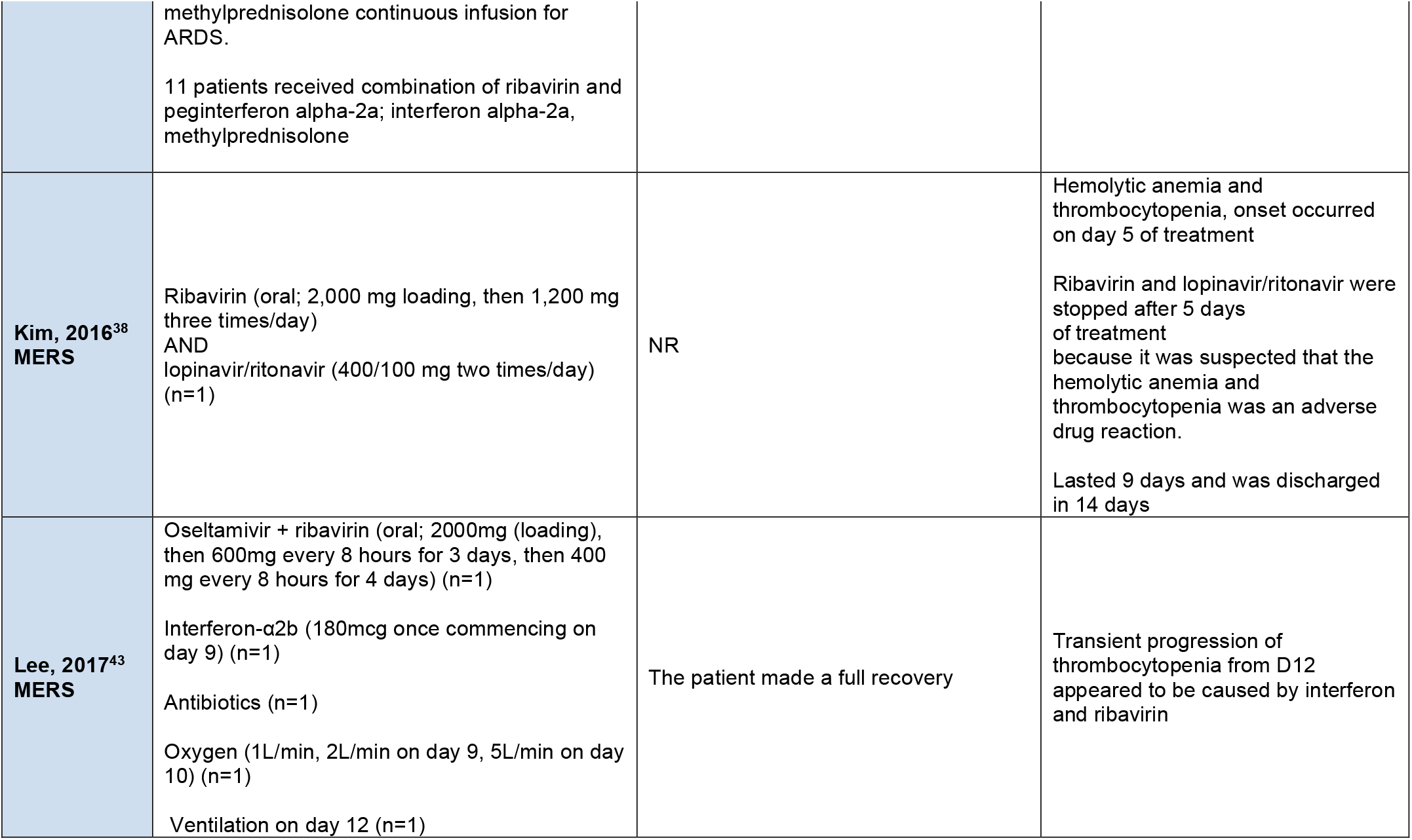
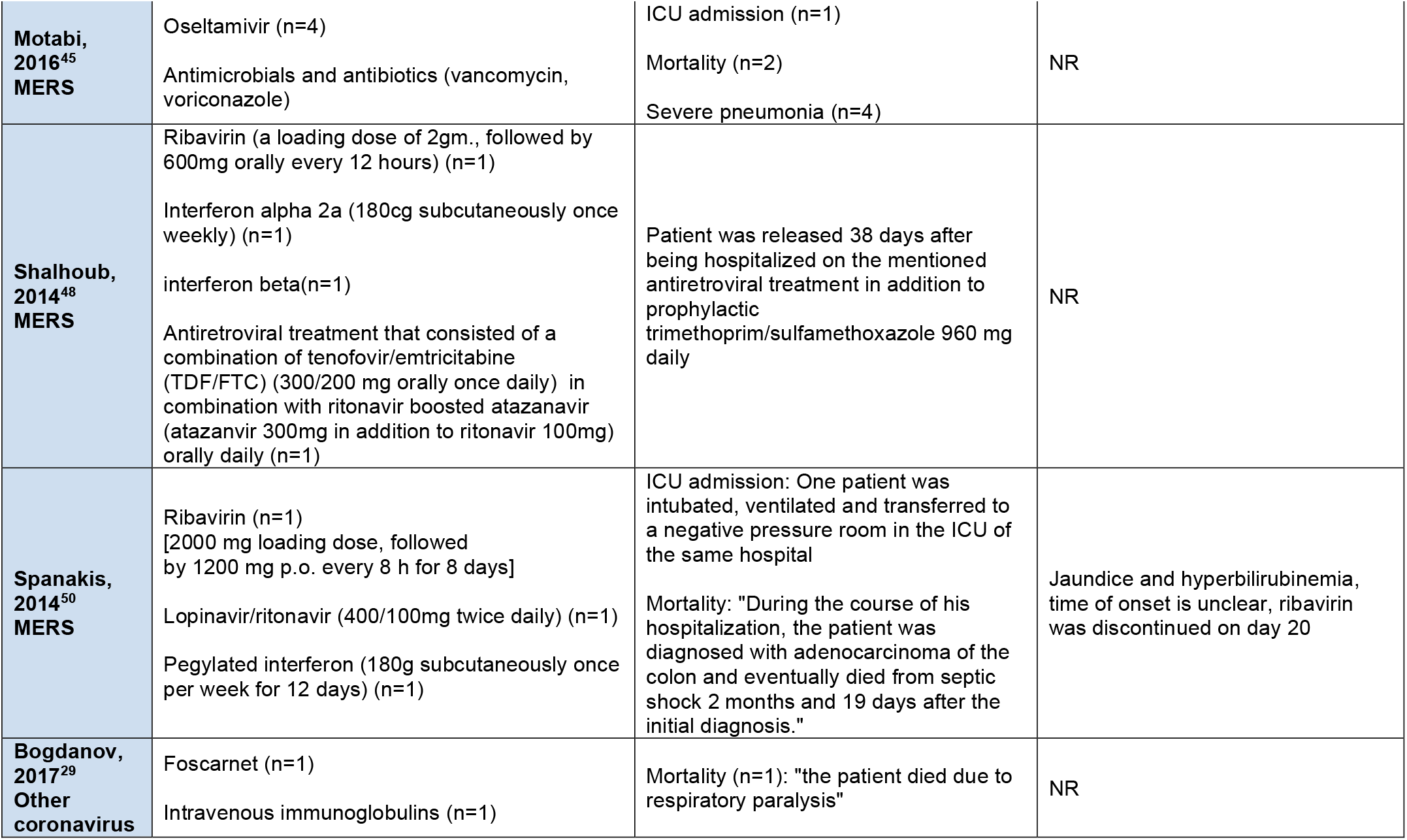
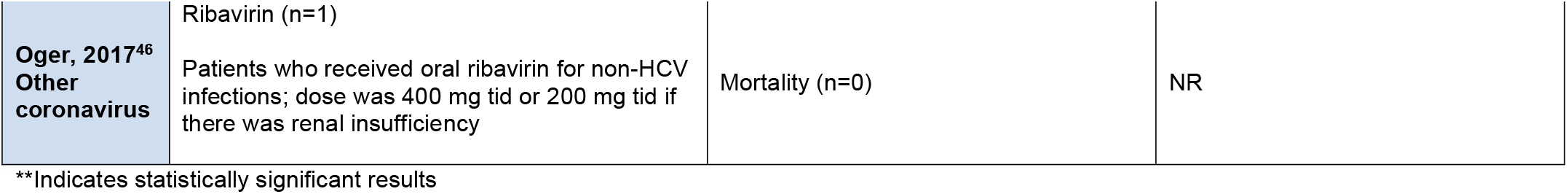

## APPENDIX 5 Quality Appraisal/Risk of Bias – Complete Results

APPENDIX 5 – Quality Appraisal/Risk of Bias – Complete Results
| Cochrane Risk of Bias Tool – Randomized Controlled Trials |  |  |  |  |  |  |  |
| --- | --- | --- | --- | --- | --- | --- | --- |
| First author | Random sequence generation | Allocation concealment | Blinding of participants and personnel | Blinding of outcome assessment | Incomplete outcome data | Selective reporting | Other bias |
| **Lee, 2004<sup>6</sup>** | Unclear risk | Unclear risk | Unclear risk | Low risk | Low risk | Unclear risk | Low risk |
| **Park, 2019<sup>7</sup>** | Unclear risk | Unclear risk | Low risk | Low risk | Low risk | Unclear risk | Low risk |
| **Zhao, 2003<sup>8</sup>** | High risk | High risk | High risk | Unclear risk | Low risk | Unclear risk | Low risk |

| Newcastle Ottawa Scale – Cohort studies |  |  |  |  |  |  |  |  |
| --- | --- | --- | --- | --- | --- | --- | --- | --- |
| Author, Year | Representativeness of the exposed cohort | Selection of the non-exposed cohort | Ascertainment of exposure | Demonstration that outcome was not present at start | Comparability of cohorts (design or analysis) | Assessment of outcome | Was follow-up long enough for outcomes to occur | Adequacy of follow up of cohorts |
| **Alkhadhairi, 2018<sup>9</sup>** | B - somewhat representative | A - same community | A - secure record | B - no | D - no description | A - independent or blind | A - yes | A - complete |
| **Arabi, 2019<sup>10</sup>** | A - truly representative | A - same community | A - secure record | B - no | A - age and other factor | A - independent or blind | A - yes | A - complete |
| **Chan, 2003<sup>11</sup>** | B - somewhat representative | A - same community | A - secure record | B - no | A - age and other factor | A - independent or blind | A - yes | B - small number lost |
| **Choi, 2016<sup>12</sup>** | B - somewhat representative | A - same community | A - secure record | B - no | D - no description | A - independent or blind | A - yes | A - complete |
| **Chu, 2004<sup>13</sup>** | B - somewhat representative | A - same community | A - secure record | B - no | A - age and other factor | A - independent or blind | A - yes | A - complete |
| **Guo, 2019<sup>14</sup>** | B - somewhat representative | A - same community | A - secure record | A - yes | D - no description | A - independent or blind | A - yes | B - small number lost |
| **Ho, 2003<sup>15</sup>** | B - somewhat representative | A - same community | A - secure record | A - yes | D - no description | A - independent or blind | A - yes | A - complete |
| **Lau, 2009<sup>16</sup>** | A - truly representative | A - same community | A - secure record | B - no | A - age and other factor | A - independent or blind | unclear | D - no description |
| **Leong, 2004<sup>17</sup>** | B - somewhat representative | A - same community | A - secure record | B - no | A - age and other factor | A - independent or blind | A - yes | A - complete |
| **Li, 2006<sup>18</sup>** | B - somewhat representative | A - same community | A - secure record | B - no | A - age and other factor | A - independent or blind | A - yes | B - small number lost |

## Notes

**Funding Statement:** This work was supported through the Drug Safety and Effectiveness Network funded by the Canadian Institutes of Health Research.

### Competing Interest Statement

The authors have declared no competing interest.

### Funding Statement

This work was supported through the Drug Safety and Effectiveness Network funded by the Canadian Institutes of Health Research.

